# Predicting future hospital antimicrobial resistance prevalence using machine learning

**DOI:** 10.1101/2023.11.30.23299234

**Authors:** Karina-Doris Vihta, Emma Pritchard, Koen B. Pouwels, Susan Hopkins, Rebecca L Guy, Katherine Henderson, Dimple Chudasama, Russell Hope, Berit Muller-Pebody, Ann Sarah Walker, David Clifton, David W. Eyre

## Abstract

**Objectives:** Predicting antimicrobial resistance (AMR), a top global health threat, nationwide at a hospital level could help target interventions. Using machine learning, we exploit historical AMR and antimicrobial usage to predict future AMR.

**Methods:** Antimicrobial use and AMR prevalence in bloodstream infections in hospitals in England were obtained per hospital group (Trust) and financial year (FY, April-March) for 22 pathogen-antibiotic combinations (FY2016-2017-FY2021-2022). XGBoost model predictions were compared to previous value taken forwards, difference between the previous two years taken forwards and linear trend forecasting (LTF). XGBoost feature importances were calculated to aid interpretability.

**Results:** Relatively limited year-to-year variability in AMR prevalence within Trust-pathogen-antibiotic combinations meant previous value taken forwards achieved a low mean absolute error (MAE). XGBoost models performed similarly, while difference between the previous two years taken forwards and LTF were consistently worse. XGBoost considerably outperformed all other methods in Trusts with a larger change in AMR prevalence from FY2020-2021 (last training year) to FY2021-2022 (held-out test set). Feature importance values indicated that complex relationships were exploited for predictions.

**Conclusion:** Year-to-year resistance has generally changed little within Trust-pathogen-antibiotic combinations. In those with larger changes, XGBoost models could improve predictions, enabling informed decisions, efficient resource allocation, and targeted interventions.

## Introduction

Antimicrobial resistance is one of the top global health threats(1). Bloodstream infections are typically one of the most serious types of infection; given their high mortality/morbidity, they are generally treated in hospital and therefore are often used for surveillance of resistance. In high-income countries any isolated pathogens will be tested for antimicrobial susceptibility against key antibiotics, while, unfortunately most low and many middle-income countries lack the laboratory capacity to test all bloodstream pathogens, if any(2). Being able to predict future antimicrobial resistance of bloodstream infections in networks of hospitals could help target interventions and allocate resources to those most at risk, with larger predicted resistance increases or absolute rates. While estimating associations between characteristics such as age, sex and probability of resistance is important at an individual level, it is not clear how a hospital should use this information. It may be simpler for hospitals to assume that their underlying populations are broadly similar from year-to-year and estimate resistance at an aggregate level. Further, empiric treatment recommendations are generally made across an entire hospital.

Antibiotic usage is a well-known driver of antibiotic resistance(3). Several studies have investigated associations, for example using Spearman’s correlation coefficients between outpatient antibiotic usage and resistance in European countries, showing countries with higher usage had higher resistance percentages(4), or using multivariate transfer functions to demonstrate positive associations between antibiotic use and resistance rates in *Pseudomonas aeruginosa* in a German hospital. The latter also allowed a decrease in resistance following usage restriction to be identified(5). Studies have generally shown increases in usage associated with quite rapid increases in resistance, while decreases in usage were associated with no changes or very delayed and more subtle decreases[6],[7]. However, exceptions have also been observed, such as increased nitrofurantoin usage leading to no changes in nitrofurantoin resistance in *E. coli* urinary tract infections, while being associated with decreased trimethoprim resistance(8). The rarity of nitrofurantoin resistance has been explained genetically by the magnitude of the distance between two genes which require inactivation(9). A recent study used distributed lag-models to estimate the relationship between relative antibiotic usage (classified as a Z-score) and antibiotic resistance at a national and international level, using 11 years of data from 26 European countries. They showed that increases in antibiotic usage Z-score was associated with an immediate and persistent increase in resistant bacteria for the 4 following years, while decreases in usage Z-score had little impact on resistance on the same time-scale; antibiotic usage of neighbouring countries also affected resistance levels(10). To our knowledge, machine learning methods have not been widely used for predicting resistance at an aggregate level such as a hospital, a network of hospitals, or a country. One study considered a feed-forward neural network with a single hidden layer, with each input neuron being a lagged time series(11); however, while this allows for nonlinearity, it models only one time series at a time(12).

In England, National Health Service hospitals are grouped into Trusts, which are organisational units serving a geographical area or a specific specialty, therefore with multiple Trusts able to serve the same geographical area. Trusts have different antibiotic usage policies and have different resistance patterns in the population they serve. Most studies so far have focused on individual pathogens, and on understanding specifically associations between antibiotic use and antibiotic resistance in individual pathogens, with some, but not all, identifying such associations. Here, we shift focus and try to predict future resistance at a Trust-level, exploiting all historical aggregate information that we have on each specific Trust, namely historical antibiotic usage for a variety of antibiotics, and historical antibiotic resistance to the pathogen-antibiotic of interest in bloodstream infections, but also resistance in other pathogen-antibiotic combinations and the complexity of these relationships. We explore whether a well-understood and typically successful machine learning model, namely XGBoost, can outperform base comparators such as carrying the last value forwards, carrying the difference between the previous two years forwards and linear trend forecasting. The hypothesis is that this type of model can exploit interactions such as decreasing use of one antibiotic leading to increasing use of another as patients still need to be treated (e.g. ciprofloxacin use declined as it was selecting for *Clostridium difficile*, and consequently amoxicillin/clavulanic acid usage increased(13)), as well as sharing of resistance mechanisms between different antibiotics and pathogens(14–16).

## Materials and Methods

National antibiotic resistance data, at a per hospital group (Trust) level, was obtained by aggregating data in the UK Health Security Agency’s (UKHSA) Second Generation Surveillance System (SGSS), containing laboratory data supplied electronically by approximately 98% of hospital microbiology laboratories in England. We studied pathogens isolated from bloodstream infections subject to mandatory surveillance (from different calendar dates, see below), and specific pathogen-antibiotic combinations, namely:

- methicillin susceptible Coagulase-positive *Staphylococcus* species (MSSA) (Apr2016-Mar2021): doxycycline/tetracyclines, erythromycin, clarithromycin, clindamycin, vancomycin
- *Escherichia coli* (Apr2016-Mar2021) and *Klebsiella* species (Apr2017-Mar2021): ciprofloxacin, third generation cephalosporins (resistance to any of cefotaxime, ceftazidime, cefpodoxime and ceftriaxone), gentamicin, carbapenems (either meropenem or imipenem; or ertapenem where meropenem and imipenem not tested), co-amoxiclav, piperacillin/tazobactam.
- *Pseudomonas aeruginosa* (Apr2017-Mar2021): ciprofloxacin, ceftazidime, gentamicin, carbapenems, piperacillin/tazobactam.

The Trust was obtained through linkage to mandatory surveillance data collected via the Healthcare-associated Infections Data Capture System. Percentages of isolates with resistance to each antibiotic were calculated per Trust per financial year (FY, i.e. April to March), to keep winter months together in the same year. There was no missing data, in that every Trust had a number of pathogens tested and resistant in each year, even if both were zero. Small numbers of isolates/month for some Trusts and key pathogen-antibiotic combinations meant monthly data had to be aggregated to years to avoid large fluctuations. Isolates tested are assumed representative of bloodstream infections in each specific Trust, as ascertainment is presumed to be high given the severity of bloodstream infections. Intermediate susceptibility results were considered susceptible following the current definition of intermediate, susceptible under increased exposure(17), and for resistance percentages to be comparable over time.

From UKHSA, we also obtained data collected by IQVIA on monthly antibiotic usage (drug, quantity, concentration) (pharmacy dispensing) at a Trust level from April 2014(18), and used defined daily doses (DDDs)(19) per antibiotic per Trust per FY to align different drugs/concentrations. We standardised antibiotic consumption to account for Trust size(20) using Trust bed occupancy data (number of day and overnight occupied beds)(21). For example, antibiotic usage of 40 DDDs amoxicillin per 100 bed-days means 40% of inpatients receive one DDD of amoxicillin every day, an estimate of the therapeutic intensity. Trust mergers were carried backwards in time, such that results are presented based on Trusts existing as distinct entities in 2021(22). In the commonly used antibiotics, data was available for all trusts across all FYs, with very few exceptions, namely, one trust missing data usage across all antibiotics in 2019-2020 and a further two in 2020-2021 (**Table S1**). In the less commonly used antibiotics, missing data was very common, although this may indicate zero usage for those years. In our models we only included the top 34 antibiotics (based on mean usage across all Trust-FYs >1%) plus ertapenem (mean usage just below 1%, but an antibiotic of interest as it is a carbapenem, the broadest spectrum antibiotic class currently in reasonably wide usage).

Our main goal was to predict future antibiotic resistance for each Trust and pathogen-antibiotic combination based on historical resistance and antibiotic usage. We fit separate models for each pathogen-antibiotic combination as the outcome, but included prior antibiotic consumption data for all antibiotics and prior resistance data for all pathogen-antibiotic combinations in each model. We also explored the predictive performance of historical usage alone. Hence, each Trust contributed a training example to each model, containing information on all annual usage and pathogen-antibiotic resistance prevalences. We explored whether a previously highly successful machine learning model with the ability to learn non-linear relationships and interactions between different features, namely XGBoost(23), could outperform base comparators. XGBoost is not designed for time series, but with appropriate feature engineering and setup can be used for time series forecasting, especially as our time series are very short. We compared XGBoost with carrying the last value forwards, carrying the difference between the previous two years forwards and linear trend forecasting (LTF) which only considers one time series from a single Trust at a time (compared to XGBoost models which consider data across all Trusts, as well as multiple time series for each Trust as features). We explored whether including previous resistance to other pathogen-antibiotic combinations and/or usage to other antibiotics in the previous 1 vs 2 vs 3 years as features improved performance vs base comparators.

We used a training-test data split based on calendar time to train models and evaluate performance. We used percentage resistance in FY2020-2021 as our outcome for our training dataset, using all data available from prior years, and in FY2021-2022 as our outcome for our test set (excluding the first year of data for each feature), excluding Trusts testing <100 isolates per year throughout the period studied and Trust-pathogen-antibiotic-FYs with ≤10 susceptbility results to avoid fluctuations due to small numbers unduly influencing results (arbitrary thresholds). 6 FYs of historical antibiotic usage were available for training (from April 2014), 4 FYs of historical resistance (from April 2016) for *E. coli* and MSSA, and 3 FYs (from April 2017) of data respectively for both *Klebsiella* sp. and *P. aeruginosa*. When exploring predictive performance with 3, 2 and 1 FY(s) historical data, we increased the size of the training dataset by considering previous years as additional outcomes. As the test dataset remained unchanged, predictive performance results were comparable. XGBoost models were fitted with both default and tuned hyperpameters. To improve generalisability, 3-fold cross-validation on the training dataset was used to tune model hyperparameters, i.e., the number of estimators, the maximum depth and the minimum child weight (see **Supplementary Methods** for full details). We also explored whether re-fitting models choosing only features with feature importance above white noise improved performance. Feature importance was captured using mean absolute SHapley Additive exPlanations (SHAP)(24) computed on the train dataset. We explored including observed values and/or means and/or differences and/or standard deviations (data not shown as performance was similar).

We chose to minimise the mean absolute error (mean of absolute difference between true and predicted value) as it is easily interpretable and less influenced by outliers than root mean squared error. XGBoost handles missing values by default, by learning at a split of a decision tree which classification of the missing value group into each split minimises the mean absolute error, and making that classification. Missing data was present in resistance because of excluding Trust-pathogen-antibiotic-FYs with <10 results (**Tables S3-S4**), and in usage for only a few Trusts in the highest usage antibiotics (**Table S1)**. When comparing performance between different models, if, for example, the previous value was missing and therefore a prediction could not be made for this model, these Trust-pathogen-antibiotic-FYs were dropped and mean absolute errors were calculated only in Trust-pathogen-antibiotic-FYs for which predictions could be made for all models being compared. To aid model interpretability, global feature importance was captured through mean absolute SHAP values, which measure the impact each feature has on the individual predictions, therefore higher values indicate more influential features. These were computed on the test set for each Trust in each individual pathogen-antibiotic combination model.

## Results

Susceptibility data were available for 138 hospital groups (Trusts) for FYs between April 2016 and March 2022 for *E. coli* and MSSA, and April 2017 and March 2022 for *Klebsiella* sp. and *P. aeruginosa*. 19 Trusts with maximum <100 tested isolates/FY across all pathogen-antibiotic combinations were excluded completely, as small sample sizes made resistance percentages highly variable from year-to-year (**Table S2**). 16/19 excluded Trusts were specialist Trusts with typically much lower rates of bloodstream infection. Trust-pathogen-antibiotic-FYs with ≤10 susceptibility results were also excluded (**Tables S3-S4**) for similar reasons (**Figure S1**).

Antibiotic resistance prevalence varied by pathogen, antibiotic, and between Trusts over the study period (**Figure 1A**). For example, within *E. coli* the median overall resistance prevalence for amoxicillin/clavulanic acid was 43%, versus 9% for piperacillin/tazobactam, but with wide interquartile ranges (IQR) (36-49% and 6-12% respectively), reflecting Trust-level variation.

**Figure 1.**
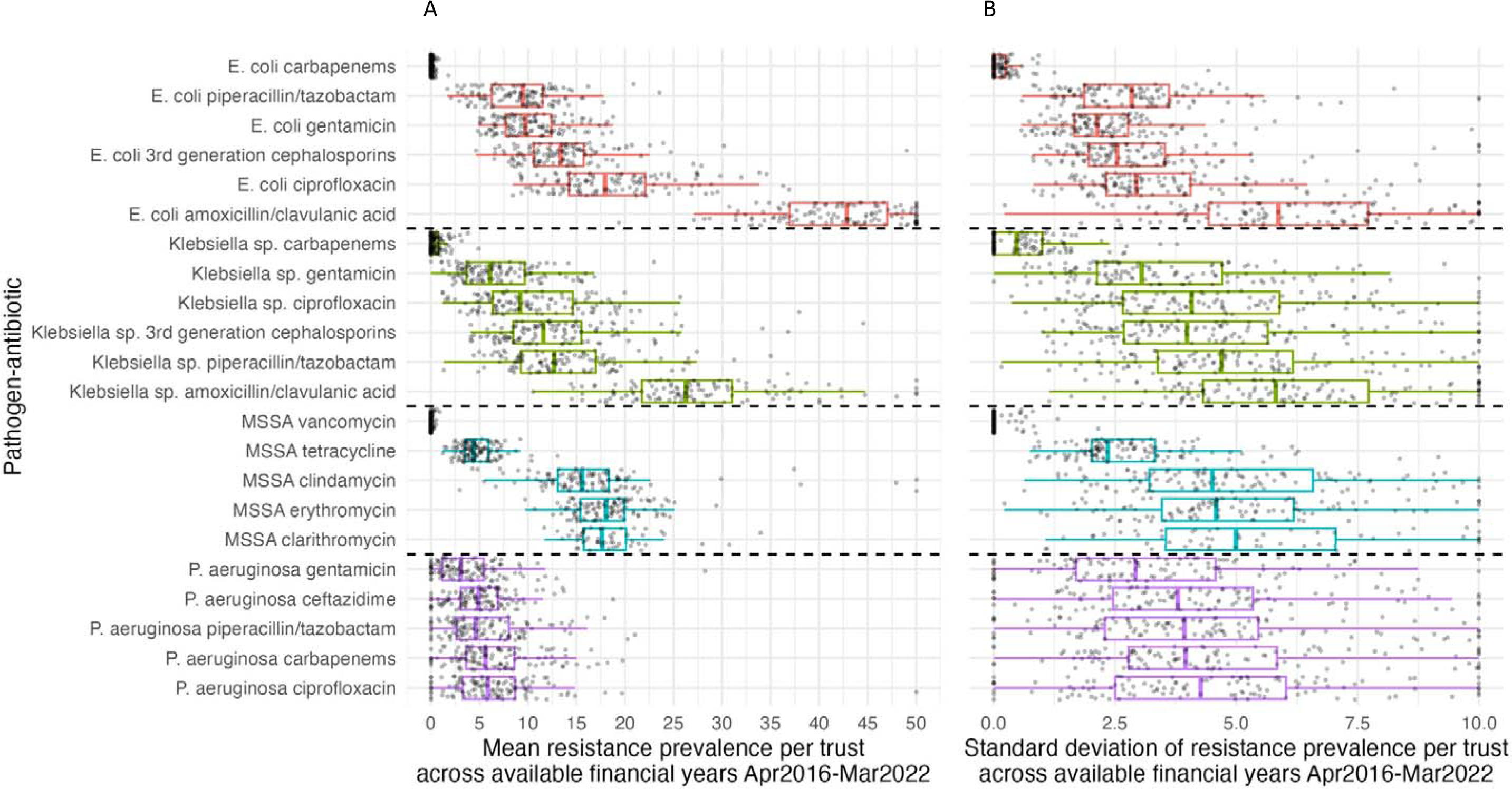
Distribution of mean resistance prevalence (A) and standard deviation (B) per Trust-pathogen-antibiotic across available financial years (Apr201 Mar2022 for E. coli and MSSA and Apr2017-Mar2022 for Klebsiella sp. and P. aeruginosa). Note: one point per Trust. Outliers outside of x-axis scale (>50 left panel, >10 right panel) were truncated.

However, there was much less variability within each Trust over time for a given pathogen-antibiotic combination, with >75% of Trusts having a standard deviation (across annual resistance prevalences) of <8% even for those pathogen-antibiotic combinations with the highest standard deviations (*E. coli*-amoxicillin/clavulanic acid and *Klebsiella* sp.-amoxicillin/clavulanic acid, **Figure 1B**). We observed uncommon outliers which may indicate data quality issues; these were not excluded from analyses as they could also represent outbreaks. Distributions of antibiotic resistance within a pathogen-antibiotic combination were broadly similar across the FYs (**Figure S2**).

Over all Trust-FYs, the median difference between current and previous resistance prevalence within each pathogen-antibiotic combination was always within ±1%; 18/22 pathogen-antibiotic combinations had median within ±0.5% (**Figure 2A**). Considering individual years, 95% of Trust-pathogen-antibiotic-FYs differed in the resistance prevalence compared with the previous year by <10% and 84% <5%. The largest absolute differences were observed for amoxicillin/clavulanic acid resistance in *Klebsiella* sp., but even there 43% of Trust-FYs had absolute differences <5%. Distributions and percentages were broadly similar over time (**Figures S3-S4**). The median LTF estimated change between FYs 2016-2017 and 2021-2022 was <2.5% for 18/22 pathogen-antibiotic combinations and <5% for the remaining 4; 82% of Trust-pathogen-antibiotic-FYs combinations had an LTF-estimated absolute change across the 6 FYs <10%, and 60% <5% (**Figure 2B**).

**Figure 2.**
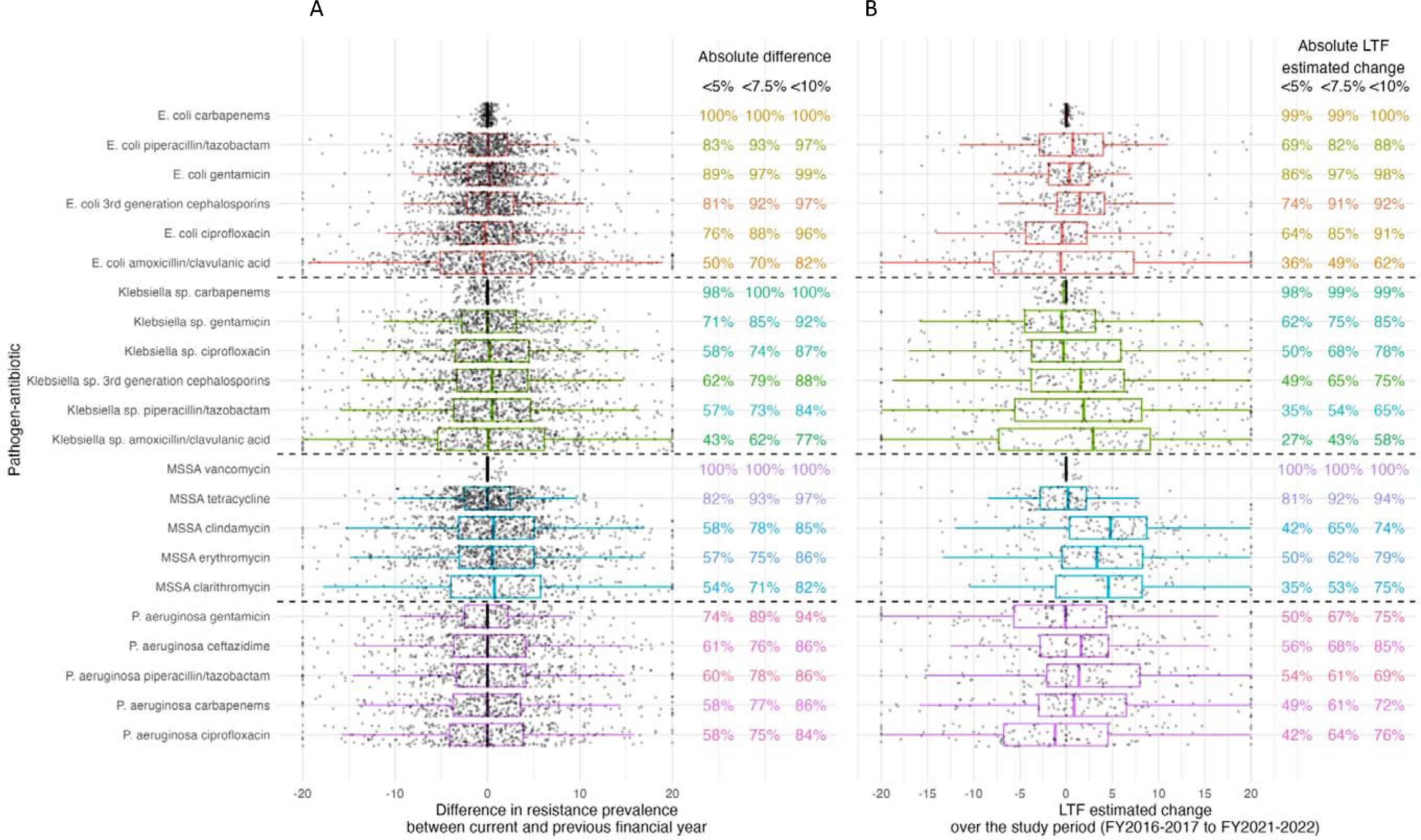
Distribution of difference in resistance prevalence between the current and previous financial year (A) and linear trend forecasting (LTF) estimat change over the study period (B), per pathogen-antibiotic combination across all Trusts and financial years. Percentages of Trust-FYs that have an absolut difference <5%, <7.5% and <10% between the current and the previous financial year are also given by pathogen-antibiotic combination (A), and an abso LTF estimated change <5%, <7.5% and <10% (B). Note: one point per Trust-year. Distribution split by financial year available in **Figure S3**. Percentage of trusts with absolute difference in resistance prevale <5%, <7.5%, and <10% split by financial year in **Figure S4**. Outliers outside of x-axis scale (absolute value >20) were truncated.

Antibiotic usage rates were available for all 119 Trusts from 2014-2015 to 2020-2021. Similarly to resistance prevalences, antibiotic usage rates varied between the different antibiotics (**Figure S5**), but with relatively little change over time for many antibiotics (**Figure S6**). Of the most commonly used antibiotics, there was a small increase in amoxicillin/clavulanic acid usage (median across Trusts 24% (IQR 15%-33%) in FY2014-2015 to 32% (22%-42%) in FY2020-2021), a decrease in trimethoprim usage (median 8% (5%-10%) to 3% (2%-5%) respectively), and a corresponding increase in sulfamethoxazole/trimethoprim (median 3% (2%-5%) to 6% (4%-9%) respectively) and in nitrofurantoin (median 2% (2%-4%) to 5% (4%-7%) respectively) (reflecting change in antibiotic recommendations for treating urinary tract infections). There was a decrease in piperacillin/tazobactam usage in FY2017-2018 to median 3% (2%-4%) (vs 5% (4%-7%) in FY2014-2015), resulting from shortages due to an explosion at a Chinese antibiotics factory(25), followed by a slow rise back to similar levels by FY2020-2021 (median 5% (3%-7%)). There was very little difference from year-to-year within a Trust, except for a few outliers, that may indicate potential data quality issues rather than true changes, potentially excepting supply interruptions and/or COVID-19 impacts (**Figures S7-S8**).

The mean absolute error from using the previous resistance prevalence taken forwards by pathogen-antibiotic-FY over the Trusts was similar over time (**Figure 3**) and approximately proportional to the mean resistance level across Trusts (**Figure S9**). Previous value taken forwards, XGBoost with default hyperparameters, and with tuned hyperparameters, as well as XGBoost models with historical antibiotic usage alone (no resistance) as features generally had a very similar performance, with no single method having the smallest mean absolute error consistently across all pathogen-antibiotic combinations (**Figure 3**, **Figure S10**). The largest differences between previous value taken forwards and XGBoost were when XGBoost outperformed previous value taken forwards, eg for *P. aeruginosa* ceftazidime (2% difference, from 4% to 6%). Taking the difference between the previous two years forwards performed the worst across all pathogen-antibiotic combinations, having the highest mean absolute error, followed by LTF. For 3 pathogen-antibiotic combinations, carbapenem resistance in *E. coli* and *Klebsiella* sp. and vancomycin resistance in MSSA, most Trusts had 0% resistance for all available FYs (**Table S5**). This was reflected in the considerably lower mean absolute error.

**Figure 3.**
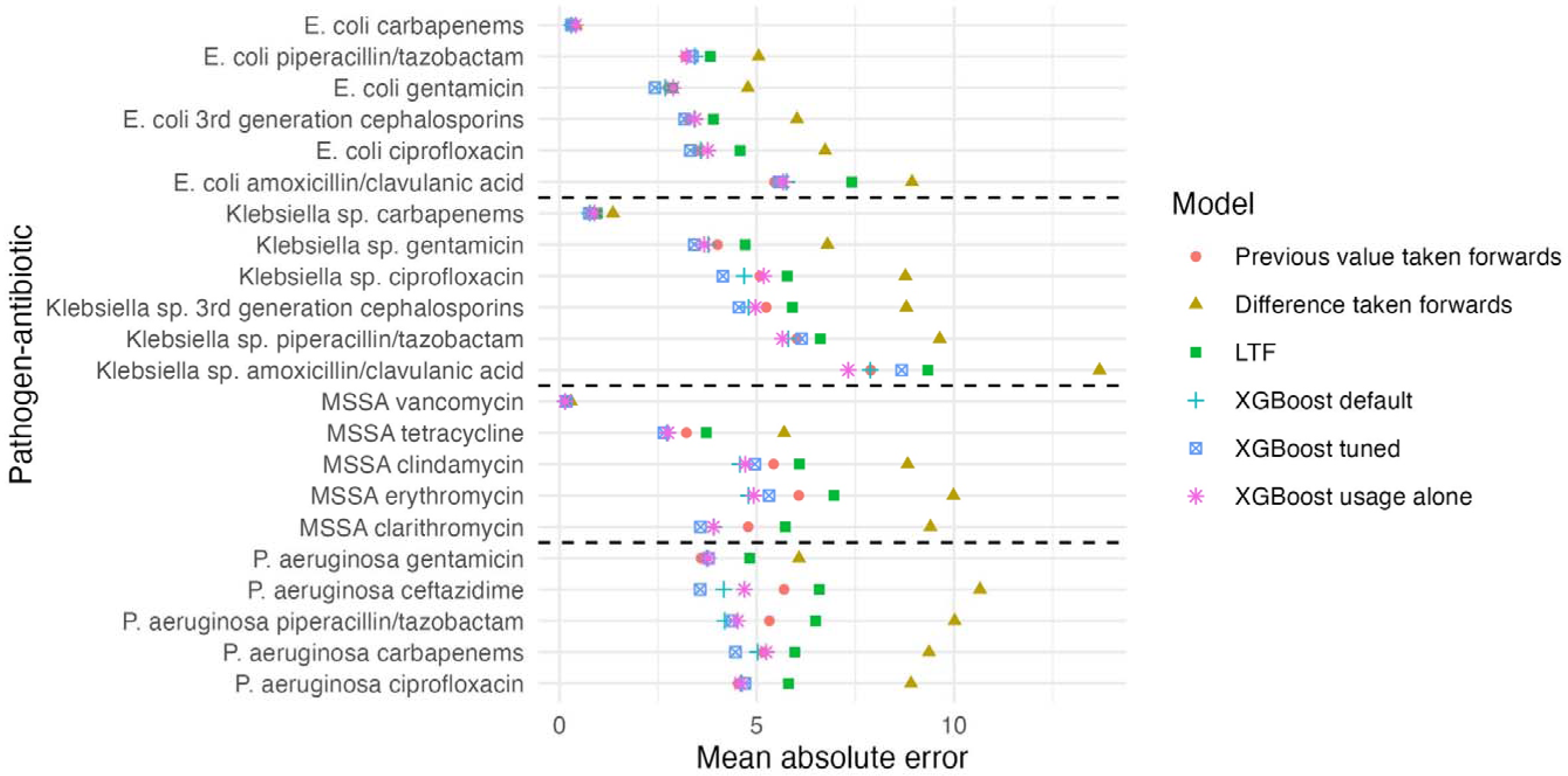
Mean absolute error for prediction on test set (resistance prevalence in FY2021-2022) for 6 different prediction models: taking the previous value forwards, taking the difference forwards, LTF, XGboost with default parameters, XGboost with tuned hyperparameters, and XGboost with previous antibiotic usage alone as input features (no information on previous resistance prevalence). Note: 70 residuals that had either missing previous value or previous difference were excluded for comparability of performance measures between the models, although XGboost also made these predictions.

XGBoost models with 3, 2 and 1 FY(s) historical data, both usage and resistance (but increased size of the training dataset by considering previous years as additional outcomes) had very similar performance (**Figure S10**). Differences in performance between XGBoost with and without feature selection where very small and neither outperformed the other across all pathogen-antibiotic combinations (**Figure S11**).

Focusing on evaluating performance in those Trusts where there was the biggest absolute difference between the resistance prevalence in FYs 2021-2022 and 2020-2021, considering an arbitrary threshold of >10% (**Figure 4**), XGBoost outperformed previous value taken forwards in all but one pathogen-antibiotic combination (*E. coli*-gentamicin). Performance gains were substantially larger in magnitude in this subgroup, while there was little to no difference in the mean absolute error in the remaining Trusts (≤10% difference). Results were similar for thresholds for the difference between resistance prevalence of 7.5% and 5%, where the outperformance by XGBoost occurred across all pathogen-antibiotic combinations including *E. coli*-gentamicin (**Figure S12**). In Trusts where the absolute difference was >10%, there were both increases and decreases from the previous resistance prevalence in 17/22 pathogen-antibiotic combinations. Performance gains in mean absolute error were observed both in Trusts with positive and negative differences between current and previous resistance prevalences (**Figure S13**).

**Figure 4.**
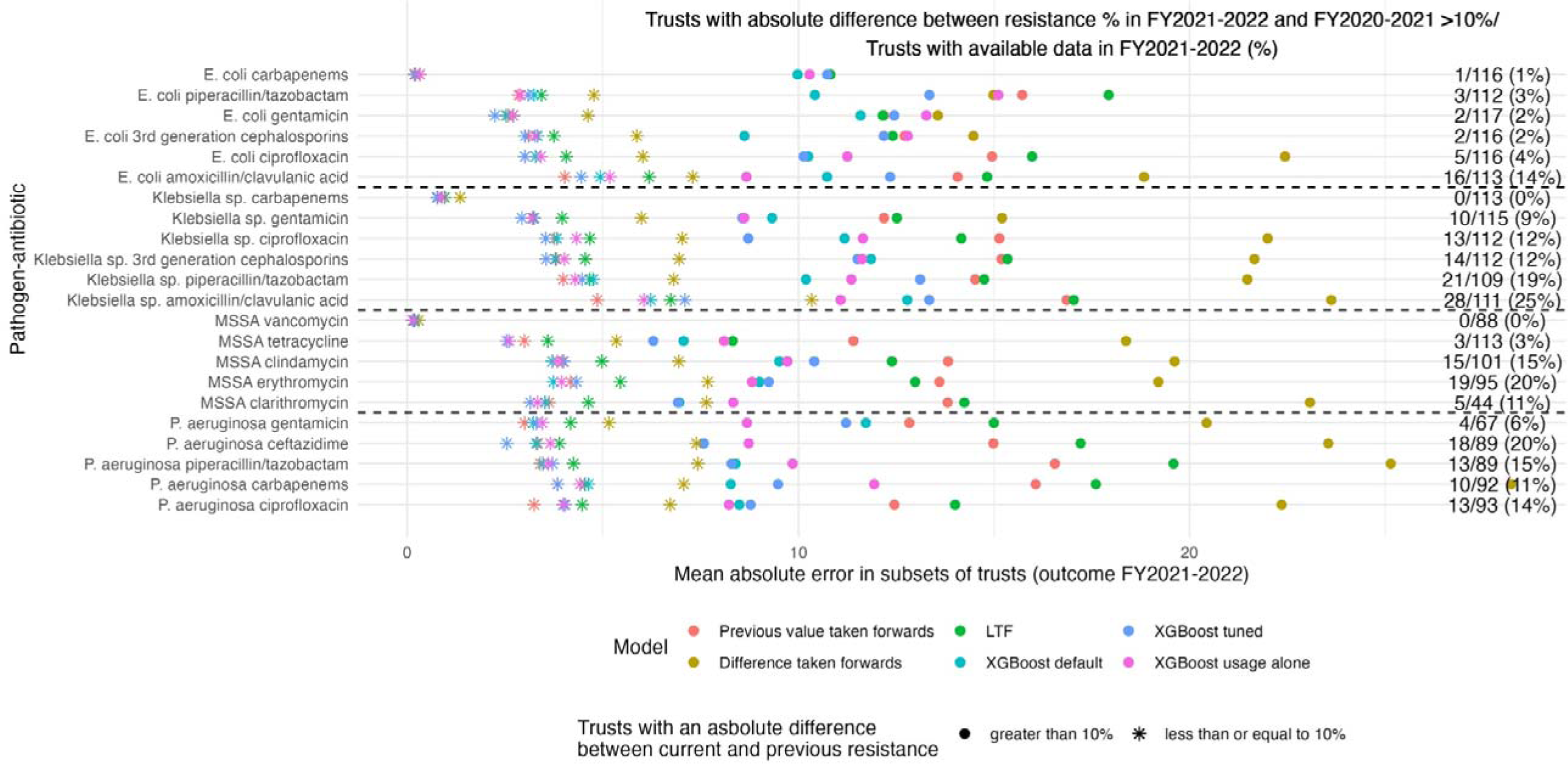
Mean absolute error for prediction on test set (resistance prevalence in FY2021-2022) for 6 different prediction models split by absolute difference between FY2021-2022 and FY2020-2021 in resistance prevalence, >10% or ≤10% Note: 70 residuals that had either missing previous value or previous difference were excluded for comparability of performance measures between the models, although XGboost also made these predictions. Results using thresholds of 7.5% and 5% are illustrated in **Figure S11**. For 3 pathogen-antibiotic combinations: E. coli carbapenems, Klebsiella sp. carbapenems and MSSA vancomycin, most Trusts had 0% resistance prevalence for all available FYs (**Table S3**).

Considering model interpretability, generally previous resistance prevalence to the same pathogen-antibiotic combination as the outcome was among the top 10 features ranked according to their mean absolute SHAP values (**Table 1**). Previous resistance prevalence to the same antibiotic but in a different pathogen, as well as usage of the same antibiotic were also generally among the top 10 features, and similarly for other antibiotics from the same class.

**Table 1.**
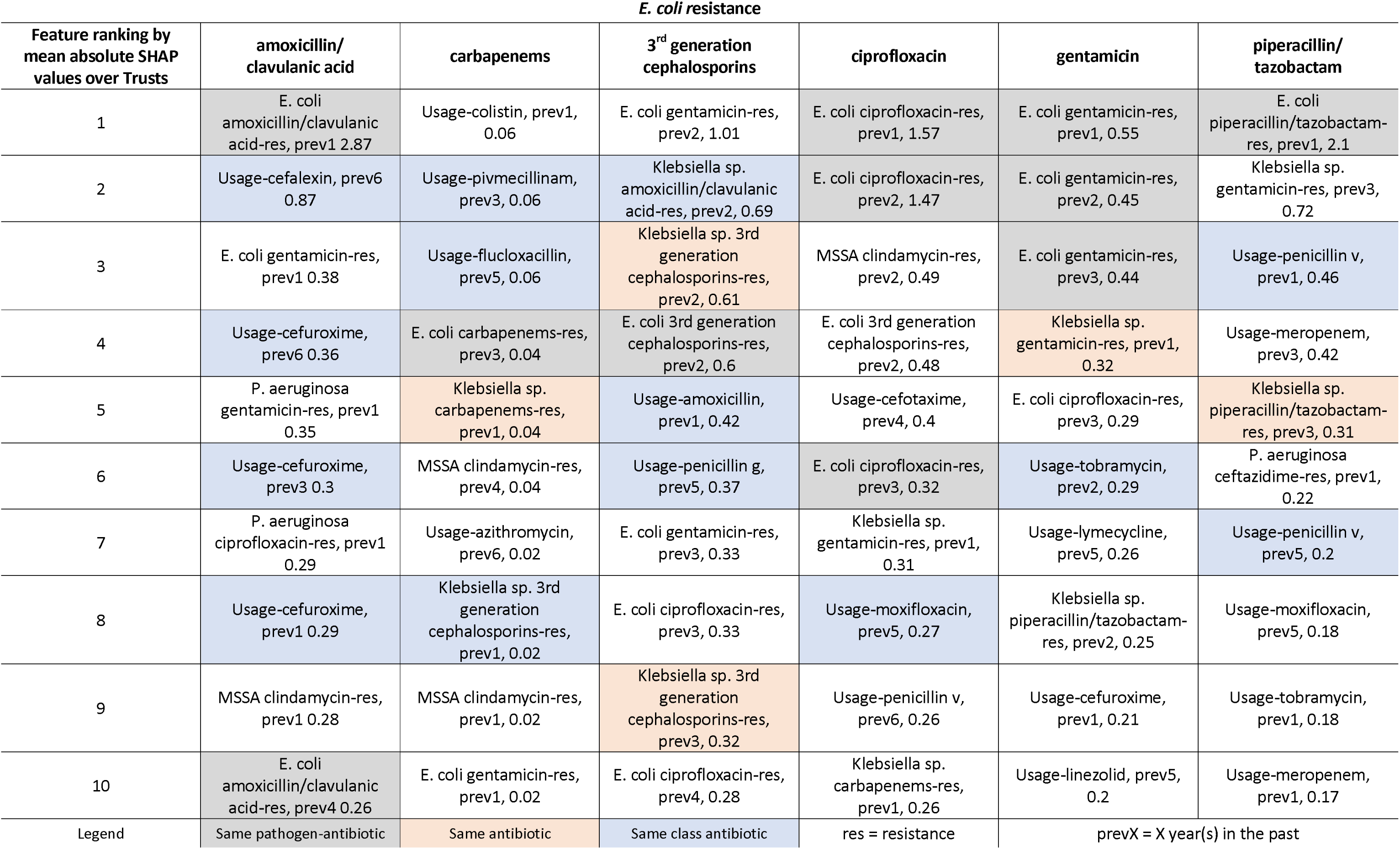
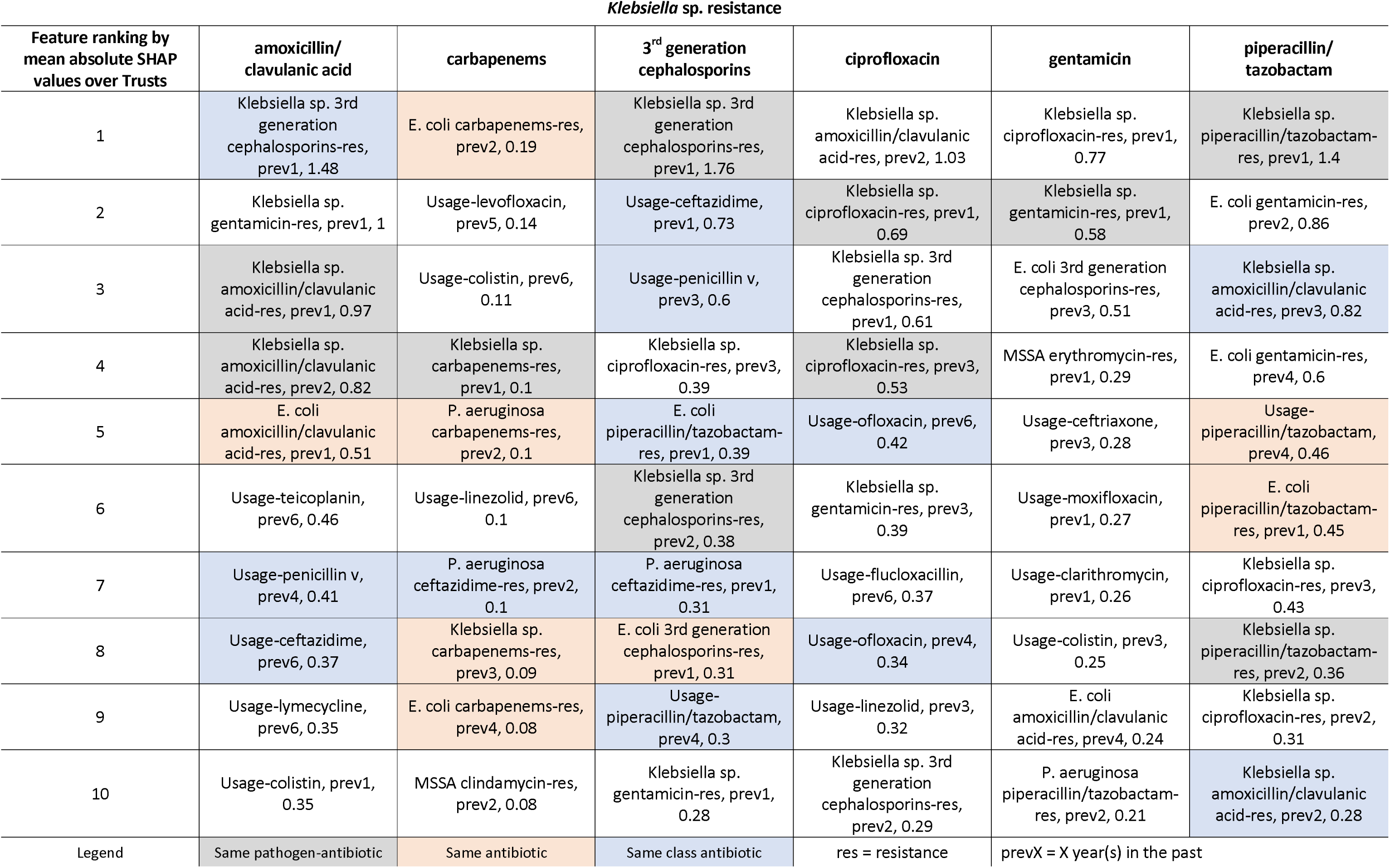
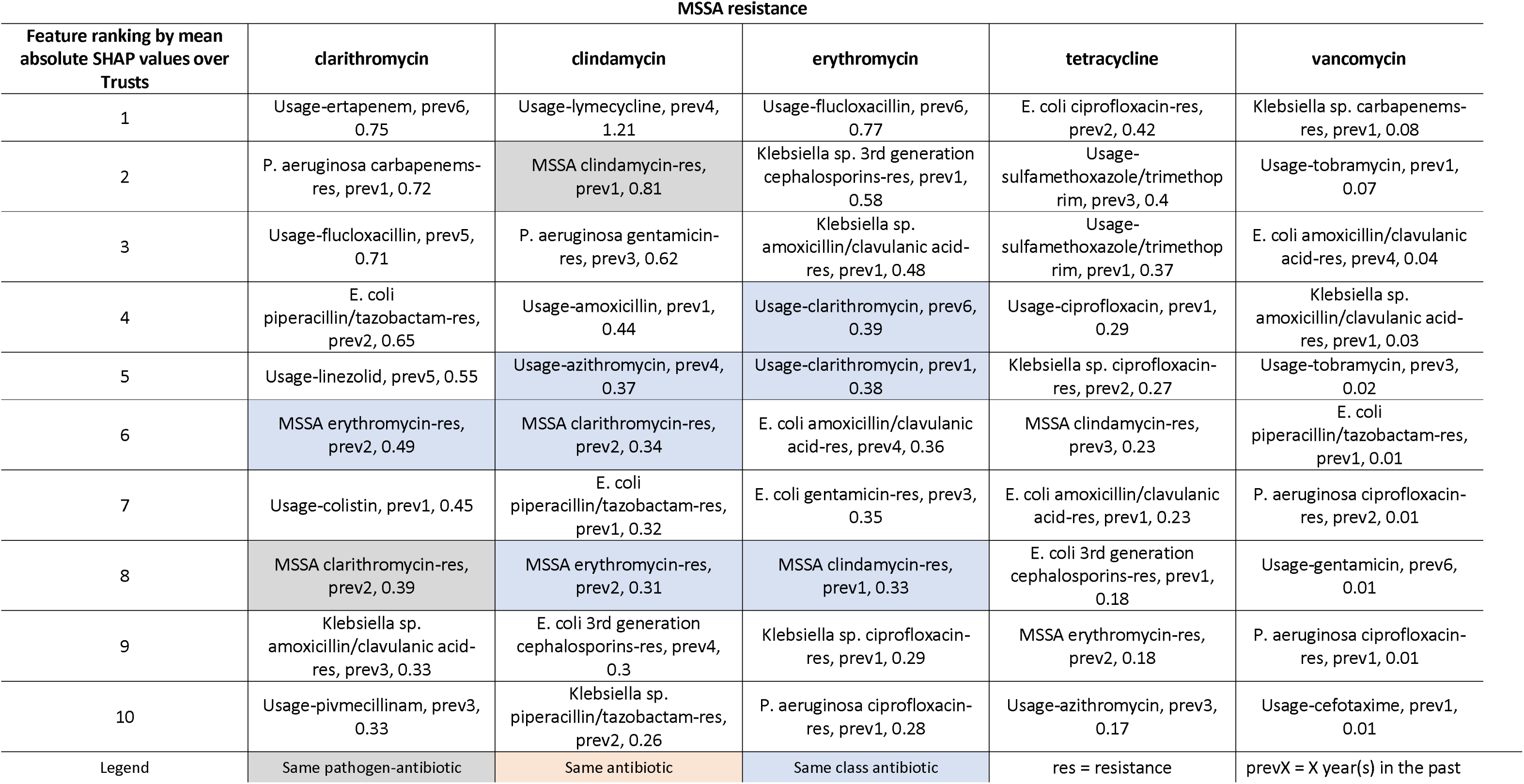
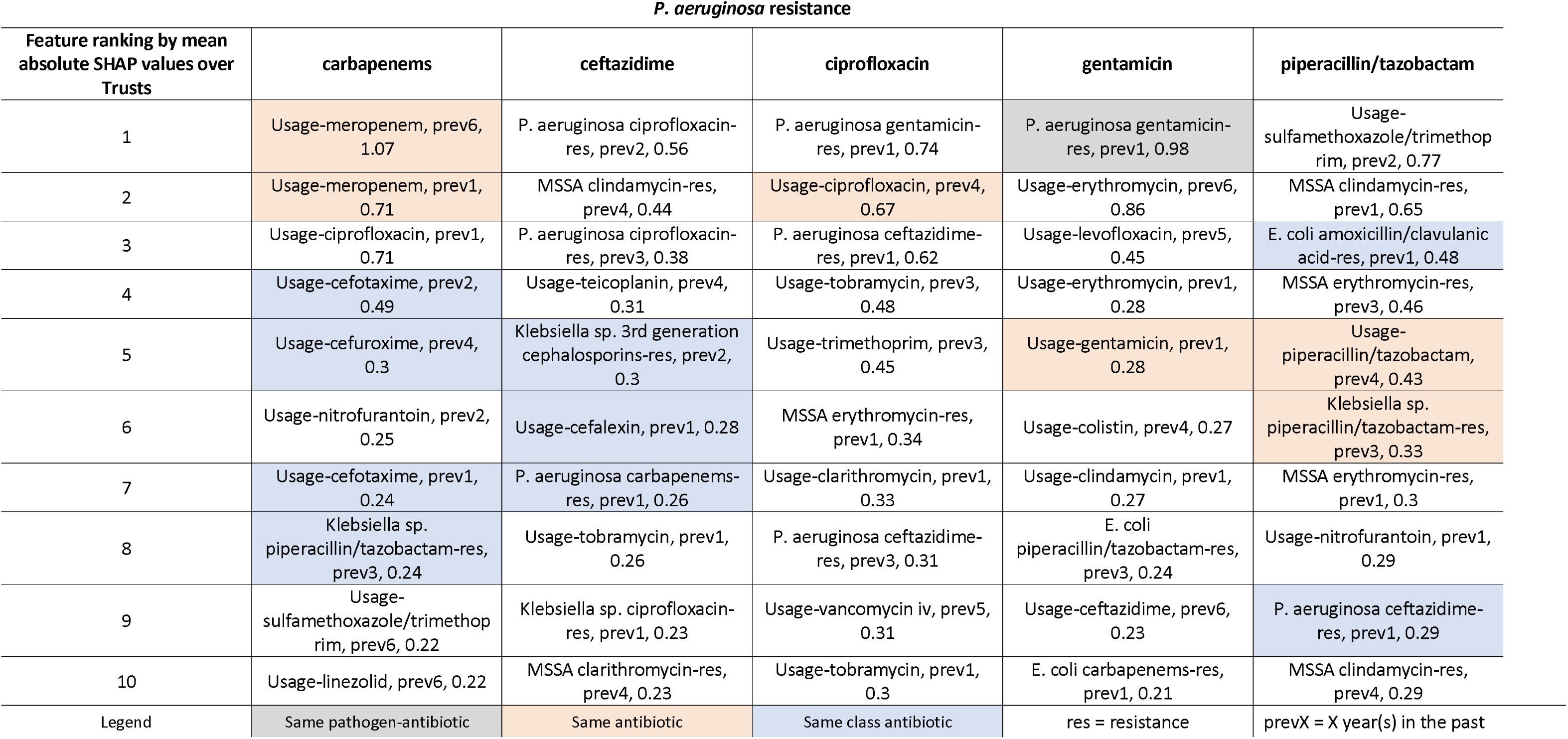
Top 10 features ranked according to mean absolute SHapley Additive exPlanations (SHAP) values for each pathogen-antibiotic combination outcome, calculated on the test dataset.

## Conclusions

While associations between antibiotic usage and antibiotic resistance are widely accepted, here we have built a model that allows us to take advantage of the complex relationship between the usage of different antibiotics, and between different resistance mechanisms being responsible for resistance to multiple antibiotics or in multiple pathogens, with the goal of predicting future resistance at an aggregate level for a hospital group. Features with the highest contributions to the prediction illustrated that such complex relationships were very likely captured and exploited by the models. One key challenge is that changes in resistance were small for many pathogen-antibiotic combinations we considered. Training the model on all Trusts, but evaluating performance in the subgroup of Trusts where changes from one financial year to the next were the largest, we achieved better predictive performance when considering the mean absolute error, without compromising predictive performance in those where the changes were minimal.

Relatively few studies have considered multiple pathogen-antibiotics simultaneously. One previous study considered forecasting quarterly resistance in *E. coli* bloodstream infections to third-generation cephalosporins, ciprofloxacin, gentamicin, and piperacillin/tazobactam per clinical commissioning group (CCG, groups of general practices) in England using data from October 2015 to October 2018, as well as annual resistance in European countries to carbapenems and fluoroquinolones in *K. pneumoniae*, *E. coli*, *P. aeruginosa*, and *Acinetobacter* spp. using data from 2012-2016(12). They compared last value taken forwards with single time series models allowing for more complexity including autoregressive integrated moving average (ARIMA), Expected-Trend-Seasonal, and a feed-forward neural network with a single hidden layer, with each input neuron being a lagged time series, as well as fitting an integrated nested Laplace approximations spatiotemporal model to all groups (CCGs or countries) and forecasting for each individual time series, while also including covariates such as antibiotic usage. They found that the median root mean square error across each pathogen-antibiotic combination was relatively small (range 0%-7%). Similarly to our study, last value taken forward outperformed the other predictors when considering aggregate performance measures for yearly European resistance data, despite the spatiotemporal model being able to capture and account for associations between antibiotic usage and resistance. At the CCG level, the more complex Expected-Trend-Seasonal model captured some seasonality and improved predictive performance, but only very slightly compared to previous value taken forwards. Traditional time series generally consider modelling one time series at a time, and while this has advantages, information on other time series or covariates can often be helpful for prediction. ARIMA is one such model, while VARIMA is an extension that considers multiple time series for forecasting. While the number of *E. coli* bloodstream infections per quarter would have also been reasonable outcome to predict for our models, we wanted to apply the same method across all pathogens under mandatory surveillance. As the numbers of isolates tested for susceptibility per quarter for all other pathogens were considerably smaller (**Figure S1**), and given the short length of our time series, we did not consider these models.

While XGBoost was not designed with time series in mind, with the right feature engineering we could use it to address our problem, providing for each Trust-pathogen-antibiotic outcome, input features comprising the historical resistance for that Trust for all available pathogen-antibiotic combinations (not just the outcome), as well as all historical antibiotic usage rates for that Trust. One strength of our analysis is our in-depth domain knowledge: standardising usage for comparison between different Trusts, antibiotics and antibiotic formulations, and financial years, by using DDDs and number of occupied beds, rather than data-agnostic standardising methods; using resistance prevalences which are appropriate to answer questions about resistance in the population at risk. We also carefully considered the changes that we could expect to be able to predict from year-to-year in the context of the distribution of resistance prevalences, allowing us to demonstrate that XGBoost models indeed achieve better predictive performance in those Trusts where there were larger changes, without impairing performance in those where changes were smaller. Taking the difference between the previous two years forwards was the worst performing model across all pathogen-antibiotic combinations, followed by LTF regression, indicating that some of the year-on-year observed differences may have been artefactual, related to the small numbers of isolates being tested and lack of representativeness of the population at risk. While LTF regression mitigated some of these fluctuations, it was either still influenced to a certain extent by the outliers and/or the linear model was not a good fit. For example, previous work found evidence for a sigmoid pattern in resistance trends, that is a fast rise following an initial period of low resistance levels, followed by a stable trend once a certain resistance percentage (below 100%) was reached(26). However, for antibiotics that have been widely used for a long period of time, our period likely only covered the stable trend, not requiring a sigmoid. Given how little year-to-year variation there was, it was difficult for average measures of performance to massively outperform previous value taken forwards, despite their ability to learn from previous resistance prevalences for all pathogen-antibiotic combinations, as well as previous antibiotic usage rates. Further, the hyperparameter tuning, feature selection, and feature engineering that we considered to improve generalisability did not improve performance (even though overfitting was reduced), with minimal decrease in mean absolute error in only some pathogen-antibiotic combinations and occasionally very small increases in some others. This is likely due to the reasonably small number of training examples which did not provide enough power to allow for learning of better hyperparameters than the default ones, which were set by the author of XGBoost based on empirical experimentation to work well on a diverse range of datasets.

One limitation of our study is the assumption that the bloodstream infections whose pathogens are tested for antibiotic susceptibility in each Trust are representative of the population being served by each Trust. For a high-income country, this may be reasonable given the severity of bloodstream infections means the vast majority of at-risk patients would have blood cultures taken, unlike low and middle-income countries where blood cultures are often only taken after empirical treatment failure(27). Another limitation is the imperfect denominator for antibiotic usage as not everyone who occupied a day or overnight bed would have received antibiotics; however, this follows World Health Organisation recommendations(21), and makes features comparable both over time and between Trusts for our prediction models. Another limitation is the data aggregation to financial years, as previous studies have shown seasonality in the usage of many antibiotics and resistance in many pathogen-antibiotic combinations in the community[6],[21]. Studies in the community rather than hospitals found the highest correlations with the antibiotics that were used most and that peaked during winter[21],[22]. Unfortunately, numbers were too small in our study for us to analyse the data quarterly across all pathogens considered; however, we did consider financial years to keep the winter months together. One alternative could have been to use smaller time periods and use both estimated resistance prevalence and some confidence limits on this (e.g. 90% CI) to represent uncertainty: however, we already had a large number of features for the number of observations. We also decided to predict resistance in FY2020-2021, despite this potentially being affected by COVID-19, given relatively limited variation in usage over time (**Figure S9**). We only tried to predict resistance one year into the future due to our short time series.

Antibiotic usage in the community, and agriculture(30), has also been shown to be associated with antimicrobial resistance. Future work could add these features into the models, although community use would need to be assigned to Trusts. While some antibiotics we considered are not used in the community, others, e.g. amoxicillin, will be very common. The small variability we observed in resistance prevalence within Trust-pathogen-antibiotic combinations could be due to the (short) length of our time series, but it could reflect a plateau if resistance had already been increasing for quite a few years before our study(26), or resistance had become balanced with antibiotic usage in most Trusts, perhaps due to antibiotic stewardship practices(31). One study used non-linear time series analysis to model relationships between antibiotic usage and resistance in five different populations in Europe for different pathogen-antibiotic combinations, as well as identify minimum usage thresholds specific to each population to guide effective antimicrobial usage, balancing effectively treating the patient with preserving the effectiveness of antibiotics(32). We note that both this study, as well as previous studies considering non-linear time series analyses to identify antibiotic usage thresholds below which no further reduction in incidence of resistance were observed, considered much longer time periods than we unfortunately had available[25],[26].

In summary, the change in resistance prevalence from year-to-year in a Trust-pathogen-antibiotic combination was generally small from FY2016-2017 onwards. However, focusing on those Trusts with larger changes, XGBoost, a machine learning model, provided better predictions of future resistance from historical antibiotic usage and historical resistance patterns in a variety of antibiotics and pathogens. Features with the highest overall contribution to predictions suggest that complex relationships were captured to achieve this performance. We therefore have a model that could be further tested and even deployed in a real-world setting to predict resistance prevalence in the next financial year, informing appropriate targeting of interventions and allocation of resources, in settings where notable changes in resistance prevalence take place.

## Author contributions

K. D. V., A. S. W., D. A. C. and D. W. E., designed the specific analysis. K. D. V. conducted the analysis. K. D. V., D. W. E. and A. S. W. drafted the manuscript. All authors contributed to interpretation of the results, revised and approved the manuscript for intellectual content, and had full access to all data analysis outputs (reports and tables) and take responsibility for their integrity and accuracy. K. D.V. is the guarantor and accepts full responsibility for the work and conduct of the study, had access to the data, controlled the decision to publish, and attests that all listed authors meet authorship criteria and that no others meeting the criteria have been omitted.

## Disclaimer

The views expressed are those of the authors and not necessarily those of the National Institute for Health Research, UK Health Security Agency or the Department of Health and Social Care. The funders/sponsors did not have any role in the design and conduct of the study; collection, management, analysis, and interpretation of the data; preparation, review, or approval of the manuscript; or decision to submit the manuscript for publication. For the purpose of Open Access, the authors have applied a Creative Commons Attribution CC BY public copyright license to any author accepted manuscript version arising from this submission.

## Acknowledgements and funding

This work was supported by the National Institute for Health Research Health Protection Research Unit (NIHR HPRU) in Healthcare Associated Infections and Antimicrobial Resistance at Oxford University in partnership with the UK Health Security Agency (NIHR200915), and the NIHR Biomedical Research Centre, Oxford. D. W. E. is a Big Data Institute Robertson Fellow. A. S. W. is an NIHR Senior Investigator. D. A. C. was supported by the Pandemic Sciences Institute at the University of Oxford; the National Institute for Health Research (NIHR) Oxford Biomedical Research Centre (BRC); an NIHR Research Professorship; a Royal Academy of Engineering Research Chair; and the InnoHK Hong Kong Centre for Centre for Cerebro-cardiovascular Engineering (COCHE). The views expressed are those of the authors and not necessarily those of the NHS, the NIHR, the Department of Health or the UK Health Security Agency.

## Data availability

A subset of the antibiotic resistance dataset is available through UKHSA’s online data service, Fingertips(35).

Information on the use of antibiotics in secondary care was obtained from IQVIA (formerly QuintilesIMS, formed from the merger of IMS Health and Quintiles). All IQVIA data used retains IQVIA Solutions UK Limited and its affiliates Copyright. All rights reserved. Use of IQVIA data for sales, marketing or any other commercial purposes is not permitted without IQVIA Solutions UK Limited’s approval, expressed by IQVIA’s Terms of Use.

