## Supplementary Material for "Predicting future hospital antimicrobial resistance prevalence using machine learning"

**Supplementary Materials and Methods**

Aggregated monthly totals for pathogen-antibiotic combinations by Trust are only available from April 2017 for *Klebsiella* spp. and *Pseudomonas aeruginosa*, and April 2016 for *E. coli* and *S. aureus*, as the Trust assignation is obtained through linkage to mandatory surveillance data. The data quality of Trust in the laboratory reported data alone has historically been poor, and while it is improving, it is not considered sufficiently accurate for prediction.

The mandatory surveillance data collection covers the *Staphylococcus* *aureus* complex which includes *S. argenteus* and *S. schweitzeri*, although *Staphylococcus* *aureus* is the predominant species within the methicillin susceptible Coagulase-positive *Staphylococcus* species (MSSA) and methicillin resistant Coagulase-positive *Staphylococcus* species (MRSA) data collections. MRSA was not considered in this study as numbers per Trust are very small even when aggregating the data to financial years, with only 694 cases in 2020/2021 being reported in England[1].

Hyperparameter tuning grid search space:

n_estimators = [10, 50, 100, 150, 200, 300, 400]

max_depth = [1, 2, 3, 4, 5, 8, 12]

min_child_weight = [3,5,7,10]

param_grid = dict(max_depth=max_depth, n_estimators=n_estimators, min_child_weight=min_child_weight)

kfold = KFold(n_splits=3, shuffle=True, random_state=7)

model = XGBRegressor(objective='reg:absoluteerror', seed=42)

grid_search = GridSearchCV(model, param_grid, scoring="neg_mean_absolute_error", n_jobs=-1, cv=kfold, verbose=1)

grid_result = grid_search.fit(X_2021_np, y_2021_np)

model_t = XGBRegressor(**grid_search.best_params_,eval_metric=mean_absolute_error,seed=42)

fit = model_t.fit(

X_2021_np,

y_2021_np,

verbose=True)

Table S1 Number of Trusts contributing data (including zero usage rate) to each financial year per antibiotic together with mean and maximum usage rate across all Trust-FYs.

|  | Number of Trusts contributing data to each financial year (N=119) | | | | | | | Mean usage rate | Max usage rate |
| --- | --- | --- | --- | --- | --- | --- | --- | --- | --- |
| Antibiotic | **2014-2015** | **2015-2016** | **2016-2017** | **2017-2018** | **2018-2019** | **2019-2020** | **2020-2021** |  |  |
| Amoxicillin/clavulanic acid | 119 | 119 | 119 | 119 | 119 | 118 | 116 | 29% | 84% |
| Flucloxacillin | 119 | 119 | 119 | 119 | 119 | 118 | 116 | 28% | 184% |
| Doxycycline | 119 | 119 | 119 | 119 | 119 | 118 | 116 | 24% | 472% |
| Clarithromycin | 119 | 119 | 119 | 119 | 119 | 118 | 116 | 17% | 58% |
| Amoxicillin | 119 | 119 | 119 | 119 | 119 | 118 | 116 | 14% | 39% |
| Metronidazole | 119 | 119 | 119 | 119 | 119 | 118 | 116 | 8% | 56% |
| Ciprofloxacin | 119 | 119 | 119 | 119 | 119 | 118 | 116 | 8% | 24% |
| Trimethoprim | 119 | 119 | 119 | 119 | 119 | 118 | 116 | 6% | 23% |
| Sulfamethoxazole/trimethoprim | 119 | 119 | 119 | 119 | 119 | 118 | 116 | 5% | 55% |
| Nitrofurantoin | 119 | 119 | 119 | 119 | 119 | 118 | 116 | 5% | 26% |
| Gentamicin | 119 | 119 | 119 | 119 | 119 | 118 | 116 | 5% | 12% |
| Piperacillin/tazobactam | 119 | 119 | 119 | 119 | 119 | 118 | 116 | 5% | 15% |
| Teicoplanin | 119 | 119 | 119 | 119 | 119 | 118 | 116 | 4% | 14% |
| Azithromycin | 119 | 119 | 119 | 119 | 119 | 118 | 116 | 4% | 46% |
| Clindamycin | 119 | 119 | 119 | 119 | 119 | 118 | 116 | 3% | 12% |
| Penicillin v | 119 | 119 | 119 | 119 | 119 | 118 | 116 | 3% | 15% |
| Levofloxacin | 114 | 115 | 118 | 118 | 119 | 118 | 116 | 2% | 22% |
| Meropenem | 119 | 119 | 119 | 119 | 119 | 118 | 116 | 2% | 9% |
| Ceftriaxone | 119 | 119 | 119 | 119 | 119 | 118 | 116 | 2% | 13% |
| Cefalexin | 119 | 119 | 119 | 119 | 119 | 118 | 116 | 2% | 15% |
| Penicillin g | 119 | 119 | 119 | 119 | 119 | 118 | 116 | 2% | 14% |
| Erythromycin | 119 | 119 | 119 | 119 | 119 | 118 | 116 | 2% | 8% |
| Cefuroxime | 119 | 118 | 118 | 118 | 118 | 117 | 116 | 2% | 13% |
| Vancomycin_iv | 119 | 119 | 119 | 119 | 119 | 118 | 116 | 1% | 8% |
| Lymecycline | 118 | 118 | 118 | 118 | 118 | 118 | 116 | 1% | 8% |
| Tobramycin | 118 | 117 | 118 | 117 | 116 | 117 | 111 | 1% | 28% |
| Cefotaxime | 119 | 119 | 119 | 119 | 119 | 118 | 116 | 1% | 8% |
| Colistin | 119 | 119 | 119 | 119 | 119 | 118 | 116 | 1% | 20% |
| Pivmecillinam | 91 | 104 | 114 | 117 | 118 | 118 | 116 | 1% | 12% |
| Ofloxacin | 113 | 114 | 114 | 115 | 115 | 109 | 109 | 1% | 9% |
| Linezolid | 119 | 119 | 119 | 119 | 119 | 118 | 116 | 1% | 3% |
| Moxifloxacin | 117 | 118 | 118 | 118 | 119 | 118 | 113 | 1% | 5% |
| Ceftazidime | 119 | 119 | 119 | 119 | 119 | 118 | 116 | 1% | 10% |
| Ertapenem | 119 | 119 | 118 | 119 | 119 | 118 | 116 | 0% | 7% |
| Cefradine | 55 | 49 | 54 | 56 | 52 | 44 | 42 | 0% | 4% |
| Daptomycin | 117 | 118 | 119 | 119 | 118 | 118 | 115 | 0% | 4% |
| Aztreonam | 108 | 109 | 110 | 105 | 108 | 110 | 108 | 0% | 8% |
| Cefaclor | 62 | 56 | 54 | 54 | 55 | 57 | 52 | 0% | 10% |
| Fusidic acid | 119 | 119 | 118 | 118 | 116 | 117 | 115 | 0% | 3% |
| Amikacin | 116 | 116 | 114 | 114 | 115 | 115 | 111 | 0% | 4% |
| Methenamine | 80 | 88 | 96 | 101 | 104 | 107 | 107 | 0% | 4% |
| Cefadroxil | 16 | 14 | 13 | 11 | 12 | 9 | 4 | 0% | 3% |
| Tigecycline | 117 | 116 | 118 | 118 | 119 | 116 | 116 | 0% | 5% |
| Temocillin | 73 | 87 | 97 | 97 | 96 | 97 | 91 | 0% | 4% |
| Oxytetracycline | 118 | 118 | 118 | 117 | 117 | 116 | 114 | 0% | 4% |
| Demeclocycline | 101 | 105 | 109 | 101 | 99 | 100 | 98 | 0% | 1% |
| Minocycline | 112 | 113 | 113 | 107 | 106 | 106 | 100 | 0% | 2% |
| Vancomycin_or | 119 | 119 | 119 | 119 | 119 | 118 | 116 | 0% | 1% |
| Cefpodoxime proxetil | 2 | 2 | 2 | 0 | 0 | 0 | 0 | 0% | 1% |
| Ampicillin/flucloxacillin | 39 | 30 | 33 | 33 | 29 | 28 | 25 | 0% | 3% |
| Neomycin | 13 | 13 | 18 | 53 | 68 | 72 | 71 | 0% | 1% |
| Fidaxomicin | 110 | 112 | 115 | 114 | 115 | 112 | 111 | 0% | 1% |
| Chloramphenicol | 117 | 117 | 118 | 117 | 116 | 116 | 114 | 0% | 3% |
| Fosfomycin | 66 | 82 | 93 | 98 | 103 | 103 | 97 | 0% | 1% |
| Cefepime | 0 | 0 | 0 | 1 | 5 | 7 | 8 | 0% | 0% |
| Cefazolin | 0 | 0 | 2 | 6 | 8 | 13 | 23 | 0% | 1% |
| Tetracycline | 98 | 98 | 99 | 101 | 102 | 98 | 94 | 0% | 0% |
| Telithromycin | 1 | 1 | 0 | 0 | 0 | 0 | 0 | 0% | 0% |
| Cefixime | 108 | 111 | 106 | 101 | 99 | 94 | 89 | 0% | 0% |
| Cilastatin/imipenem | 65 | 63 | 65 | 69 | 71 | 69 | 63 | 0% | 1% |
| Ceftolozane/tazobactam | 5 | 22 | 41 | 55 | 68 | 76 | 68 | 0% | 0% |
| Avibactam/ceftazidime | 0 | 0 | 30 | 69 | 89 | 97 | 100 | 0% | 0% |
| Tedizolid | 3 | 12 | 18 | 20 | 16 | 18 | 16 | 0% | 0% |
| Cefoxitin | 0 | 0 | 15 | 29 | 31 | 27 | 26 | 0% | 0% |
| Spiramycin | 0 | 0 | 0 | 9 | 16 | 13 | 7 | 0% | 0% |
| Ceftobiprole medocaril | 0 | 8 | 10 | 10 | 10 | 10 | 7 | 0% | 0% |
| Tinidazole | 91 | 86 | 82 | 80 | 87 | 81 | 69 | 0% | 0% |
| Ceftaroline fosamil | 19 | 18 | 23 | 27 | 21 | 27 | 28 | 0% | 0% |
| Norfloxacin | 28 | 13 | 4 | 5 | 4 | 3 | 1 | 0% | 0% |
| Meropenem/vaborbactam | 0 | 0 | 0 | 0 | 2 | 4 | 17 | 0% | 0% |
| Ampicillin | 36 | 28 | 29 | 23 | 20 | 18 | 19 | 0% | 0% |
| Dalfopristin/quinupristin | 7 | 5 | 5 | 3 | 0 | 0 | 0 | 0% | 0% |
| Doripenem | 2 | 2 | 0 | 0 | 0 | 0 | 0 | 0% | 0% |
| Cefalotin | 0 | 0 | 0 | 0 | 0 | 1 | 1 | 0% | 0% |
| Cilastatin/imipenem/relebactam | 0 | 0 | 0 | 0 | 0 | 1 | 6 | 0% | 0% |
| Piperacillin | 1 | 2 | 2 | 2 | 1 | 1 | 1 | 0% | 0% |
| Cefiderocol | 0 | 0 | 0 | 0 | 0 | 7 | 7 | 0% | 0% |
| Delafloxacin | 0 | 0 | 0 | 0 | 0 | 0 | 3 | 0% | 0% |
| Telavancin | 2 | 2 | 0 | 1 | 1 | 0 | 0 | 0% | 0% |

Table S2 Trusts with <100 isolates tested per financial year across all the pathogen-antibiotic combinations excluded from analyses

| Trust | Maximum number of tests  per pathogen-antibiotic per financial year | Trust type |
| --- | --- | --- |
| MOORFIELDS EYE HOSPITAL NHS FOUNDATION TRUST | 0 | ACUTE - SPECIALIST |
| QUEEN VICTORIA HOSPITAL NHS FOUNDATION TRUST | 0 | ACUTE - SPECIALIST |
| AIREDALE NHS FOUNDATION TRUST | 1 | ACUTE - SMALL |
| NORTH TEES AND HARTLEPOOL NHS FOUNDATION TRUST | 1 | ACUTE - MEDIUM |
| THE ROYAL ORTHOPAEDIC HOSPITAL NHS FOUNDATION TRUST | 3 | ACUTE - SPECIALIST |
| ROYAL NATIONAL ORTHOPAEDIC HOSPITAL NHS TRUST | 5 | ACUTE - SPECIALIST |
| THE ROBERT JONES AND AGNES HUNT ORTHOPAEDIC HOSPITAL NHS FOUNDATION TRUST | 7 | ACUTE - SPECIALIST |
| LIVERPOOL HEART AND CHEST HOSPITAL NHS FOUNDATION TRUST | 18 | ACUTE - SPECIALIST |
| THE WALTON CENTRE NHS FOUNDATION TRUST | 19 | ACUTE - SPECIALIST |
| SHEFFIELD CHILDREN'S NHS FOUNDATION TRUST | 21 | ACUTE - SPECIALIST |
| ROYAL PAPWORTH HOSPITAL NHS FOUNDATION TRUST | 22 | ACUTE - SPECIALIST |
| LIVERPOOL WOMEN'S NHS FOUNDATION TRUST | 23 | ACUTE - SPECIALIST |
| ALDER HEY CHILDREN'S NHS FOUNDATION TRUST | 24 | ACUTE - SPECIALIST |
| THE CLATTERBRIDGE CANCER CENTRE NHS FOUNDATION TRUST | 27 | ACUTE - SPECIALIST |
| GREAT ORMOND STREET HOSPITAL FOR CHILDREN NHS FOUNDATION TRUST | 32 | ACUTE - SPECIALIST |
| BIRMINGHAM WOMEN'S AND CHILDREN'S NHS FOUNDATION TRUST | 40 | ACUTE - SPECIALIST |
| THE ROYAL MARSDEN NHS FOUNDATION TRUST | 63 | ACUTE - SPECIALIST |
| THE CHRISTIE NHS FOUNDATION TRUST | 91 | ACUTE - SPECIALIST |
| DORSET COUNTY HOSPITAL NHS FOUNDATION TRUST | 97 | ACUTE - SMALL |

Table S3 Number of Trust-pathogen-antibiotic-FYs that were excluded due to <=10 isolates tested by pathogen-antibiotic combination and the number of Trusts from which these FYs came

| Pathogen-antibiotic combination | Number of financial years excluded  (N=5/6 FYs*119) | Number of trusts with excluded financial years (N=119) |
| --- | --- | --- |
| E.coli 3rd generation cephalosporins | 18/714 (3%) | 7 |
| E. coli amoxicillin/clavulanic acid | 27/714 (4%) | 9 |
| E. coli carbapenems | 15/714 (2%) | 6 |
| E. coli ciprofloxacin | 20/714 (3%) | 7 |
| E. coli gentamicin | 14/714 (2%) | 6 |
| E. coli piperacillin/tazobactam | 39/714 (5%) | 13 |
| Klebsiella sp. 3rd generation cephalosporins | 22/595 (4%) | 11 |
| Klebsiella sp. amoxicillin/clavulanic acid | 27/595 (5%) | 11 |
| Klebsiella sp. carbapenems | 17/595 (3%) | 9 |
| Klebsiella sp. ciprofloxacin | 22/595 (4%) | 10 |
| Klebsiella sp. gentamicin | 14/595 (2%) | 8 |
| Klebsiella sp. piperacillin/tazobactam | 37/595 (6%) | 16 |
| MSSA clarithromycin | 437/714 (61%) | 79 |
| MSSA clindamycin | 95/714 (13%) | 30 |
| MSSA erythromycin | 120/714 (17%) | 28 |
| MSSA tetracycline | 29/714 (4%) | 15 |
| MSSA vancomycin | 144/714 (20%) | 43 |
| P. aeruginosa carbapenems | 94/595 (16%) | 40 |
| P. aeruginosa ceftazidime | 94/595 (16%) | 39 |
| P. aeruginosa ciprofloxacin | 89/595 (15%) | 39 |
| P. aeruginosa gentamicin | 123/595 (21%) | 60 |
| P. aeruginosa piperacillin/tazobactam | 107/595 (18%) | 44 |

Table S4 Number of financial years that each Trust had >10 tested isolates, and hence were included in analyses, for each pathogen-antibiotic combination.

|  | Number of financial years contributing data | | | | | | Number of Trusts with any data |
| --- | --- | --- | --- | --- | --- | --- | --- |
| Pathogen-antibiotic | **1** | **2** | **3** | **4** | **5** | **6** |  |
| E. coli amoxicillin/clavulanic acid | 1 | 1 | 3 | 0 | 3 | 110 | 118 |
| E. coli carbapenems | 1 | 1 | 1 | 0 | 3 | 113 | 119 |
| E. coli 3^rd^ generation cephalosporins | 1 | 0 | 1 | 0 | 4 | 112 | 118 |
| E. coli ciprofloxacin | 1 | 0 | 2 | 0 | 3 | 112 | 118 |
| E. coli gentamicin | 1 | 0 | 2 | 0 | 3 | 113 | 119 |
| E. coli piperacillin/tazobactam | 1 | 0 | 5 | 2 | 3 | 106 | 117 |
| Klebsiella sp. amoxicillin/clavulanic acid | 2 | 0 | 2 | 5 | 108 | / | 117 |
| Klebsiella sp. carbapenems | 0 | 1 | 2 | 5 | 110 | / | 118 |
| Klebsiella sp. 3^rd^ generation cephalosporins | 0 | 0 | 3 | 6 | 108 | / | 117 |
| Klebsiella sp. ciprofloxacin | 0 | 1 | 2 | 5 | 109 | / | 117 |
| Klebsiella sp. gentamicin | 0 | 0 | 2 | 5 | 111 | / | 118 |
| Klebsiella sp. piperacillin/tazobactam | 0 | 3 | 3 | 7 | 103 | / | 116 |
| MSSA clarithromycin | 3 | 5 | 2 | 2 | 2 | 40 | 54 |
| MSSA clindamycin | 2 | 4 | 3 | 4 | 10 | 89 | 112 |
| MSSA erythromycin | 0 | 3 | 4 | 0 | 6 | 91 | 104 |
| MSSA tetracycline | 1 | 0 | 4 | 2 | 8 | 104 | 119 |
| MSSA vancomycin | 3 | 4 | 9 | 4 | 12 | 76 | 108 |
| P. aeruginosa carbapenems | 5 | 7 | 5 | 18 | 79 | / | 114 |
| P. aeruginosa ceftazidime | 5 | 11 | 6 | 14 | 80 | / | 116 |
| P. aeruginosa ciprofloxacin | 3 | 12 | 5 | 16 | 80 | / | 116 |
| P. aeruginosa gentamicin | 4 | 9 | 17 | 26 | 59 | / | 115 |
| P. aeruginosa piperacillin/tazobactam | 3 | 14 | 6 | 16 | 75 | / | 114 |

Note: For *Klebsiella* sp. and *P. aeruginosa* a maximum of 5 FYs were available (indicated through a /).

Table S5 Number of Trusts with 0% resistance for all observed financial years for each specific pathogen-antibiotic combination. The pathogen-antibiotic combinations that are missing had 0 Trusts with 0% resistance throughout.

| Pathogen-antibiotic | Number of Trusts with resistance 0% throughout  up to 2020-2021 (N=119) | Number of Trusts with resistance 0% throughout  up to 2021-2022 (N=119) |
| --- | --- | --- |
| E. coli carbapenems | **69** | **62** |
| Klebsiella sp. carbapenems | **56** | **53** |
| Klebsiella sp. ciprofloxacin | 1 | 0 |
| Klebsiella sp. gentamicin | 2 | 1 |
| MSSA tetracycline | 1 | 0 |
| MSSA vancomycin | **93** | **91** |
| P. aeruginosa carbapenems | 10 | 7 |
| P. aeruginosa ceftazidime | 12 | 9 |
| P. aeruginosa ciprofloxacin | 13 | 11 |
| P. aeruginosa gentamicin | 19 | 16 |
| P. aeruginosa piperacillin/tazobactam | 15 | 10 |

Figure S1 Difference in resistance prevalence between current and previous year by number of tests per year for the specific Trust-pathogen-antibiotic combinations.

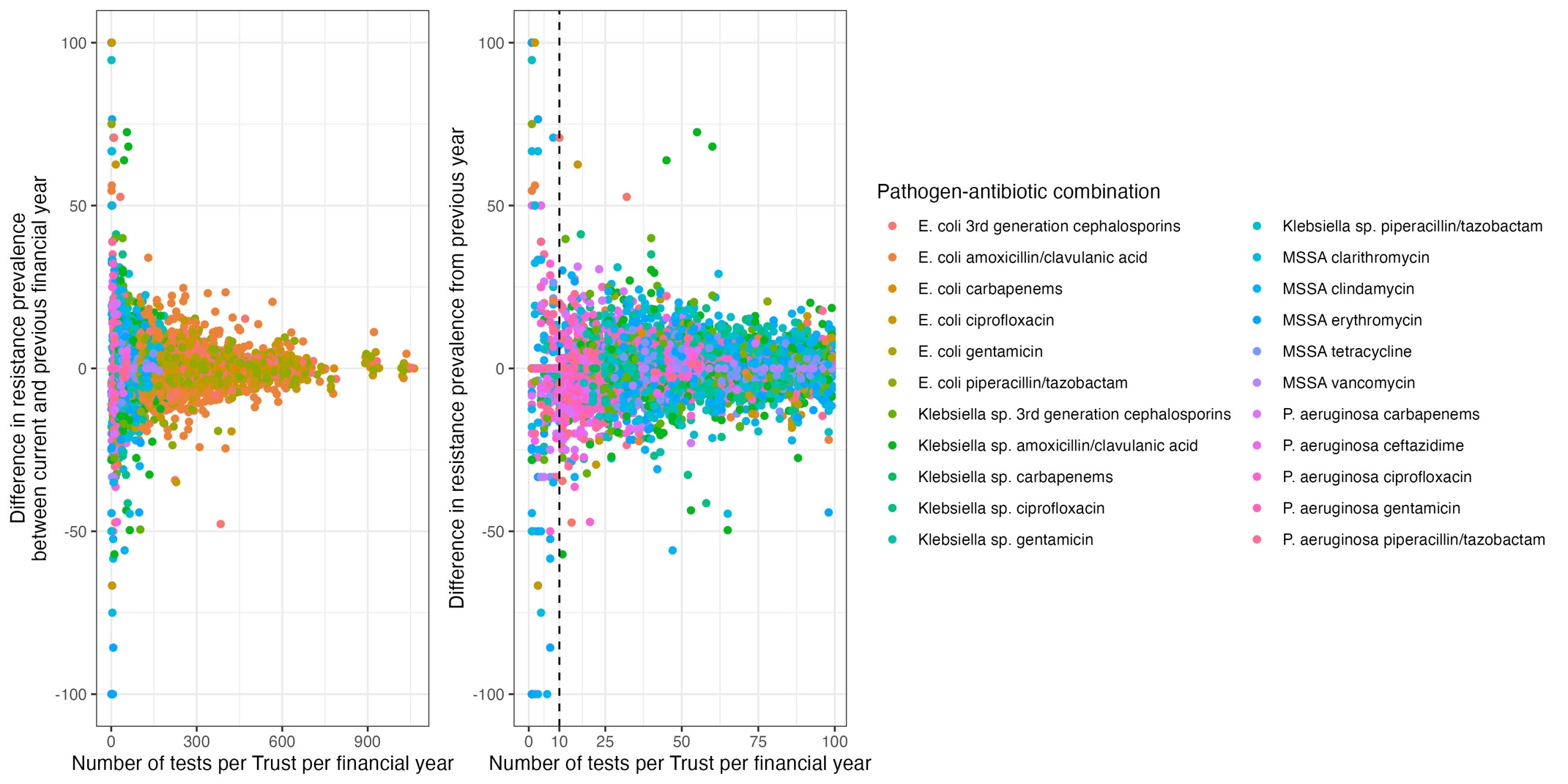

Note: one point per Trust-pathogen-antibiotic-financial year. Right-hand panel is a subset of the left-hand panel with the vertical black dashed line at the arbitrary threshold for inclusion (x=10).

Figure S2 Distribution of resistance prevalence per pathogen-antibiotic across Trusts and financial years.

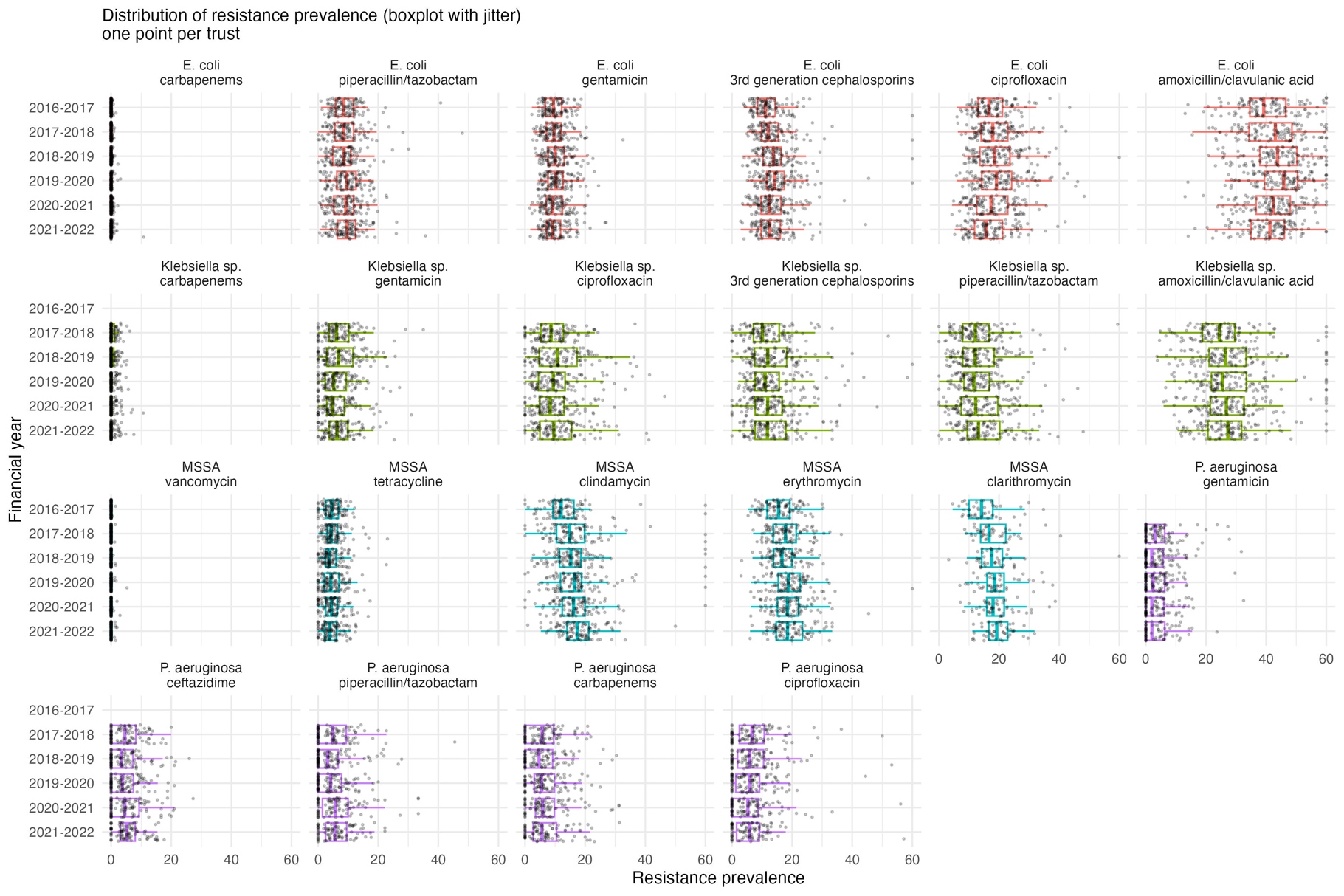

Note: one point per Trust. Outliers outside of x-axis scale (absolute value >10) were truncated.

Figure S3 Distribution of difference between the current and previous financial year resistance prevalence, per pathogen-antibiotic across Trusts and financial years.

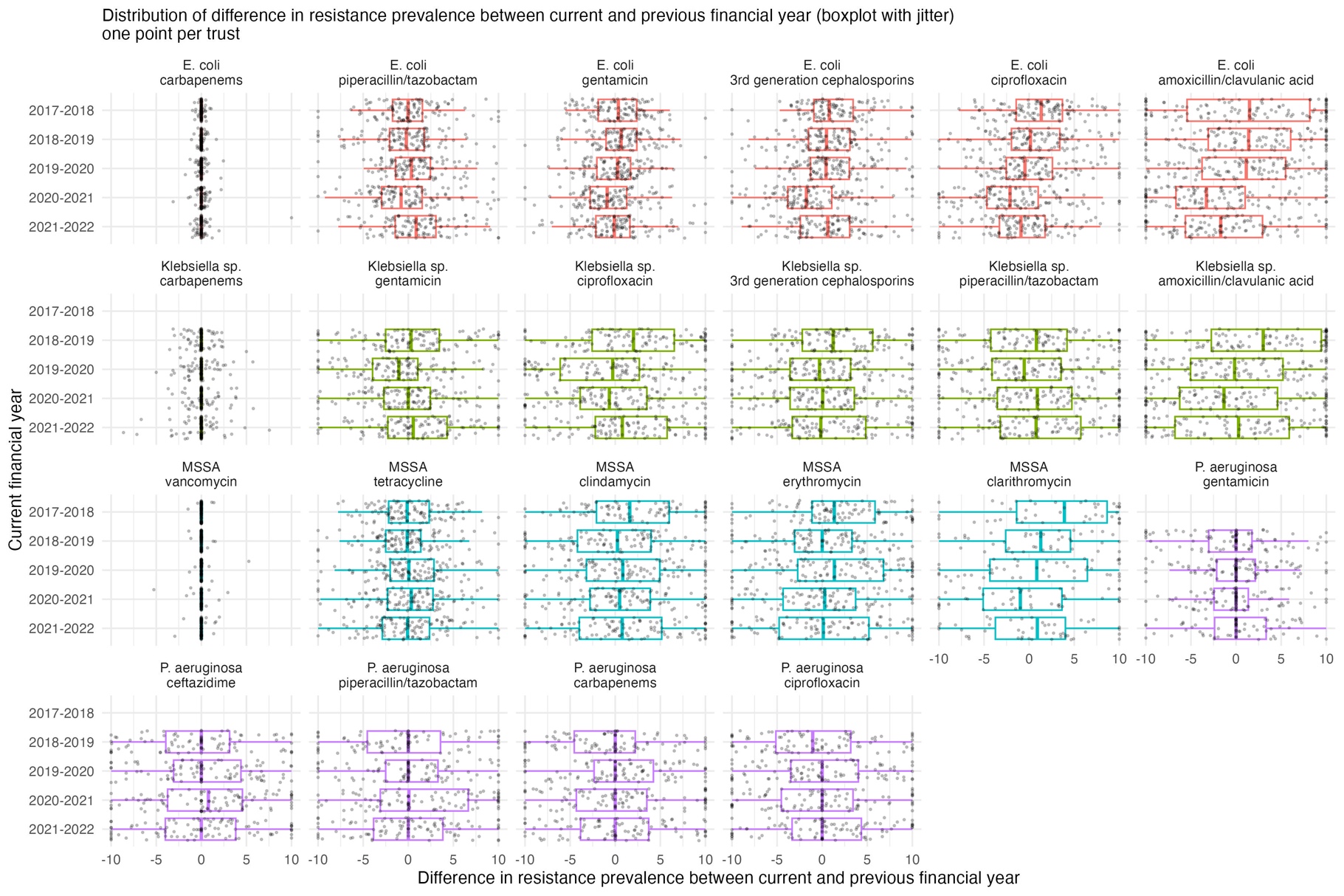

Note: one point per Trust. Outliers outside of x-axis scale (absolute value >10) were truncated.

Figure S4 Percentage of Trusts with absolute difference between the current and previous financial year resistance prevalence <5%, <7.5%, and <10% per pathogen-antibiotic-FY.

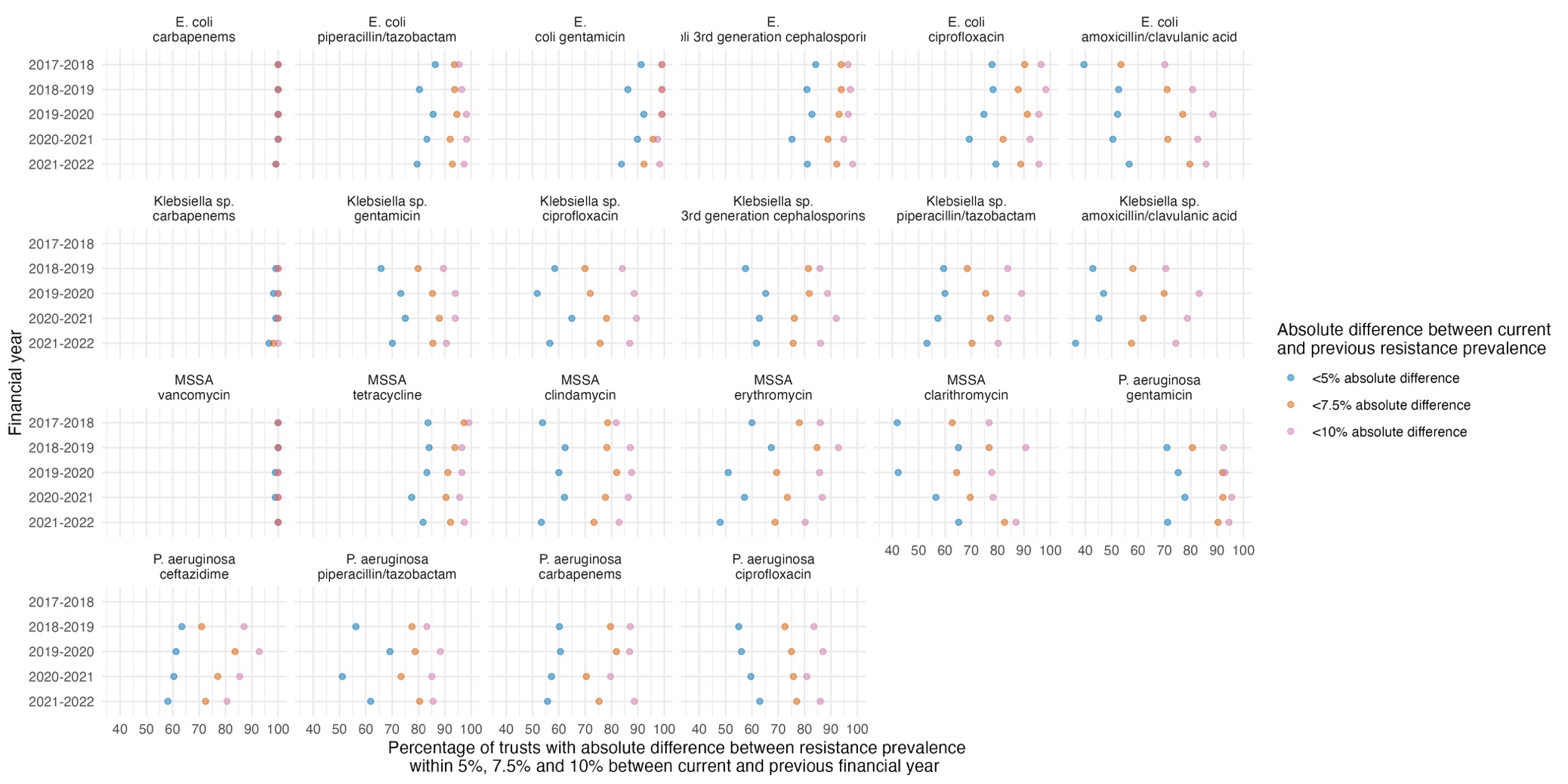

Figure S5 Distribution of mean (A) and standard deviation (B) of antibiotic usage rate per antibiotic per Trust across available financial years.

(A)

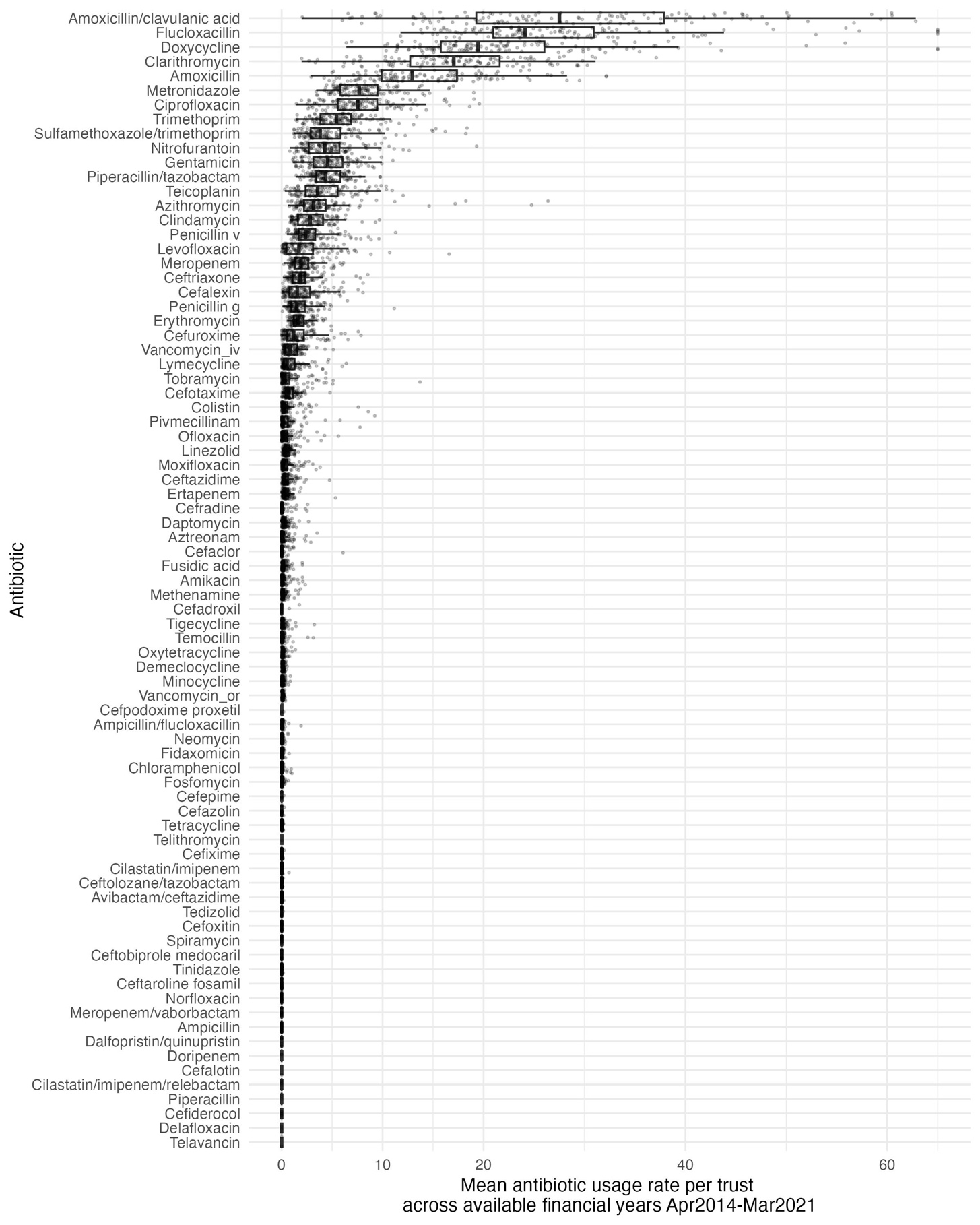

(B)

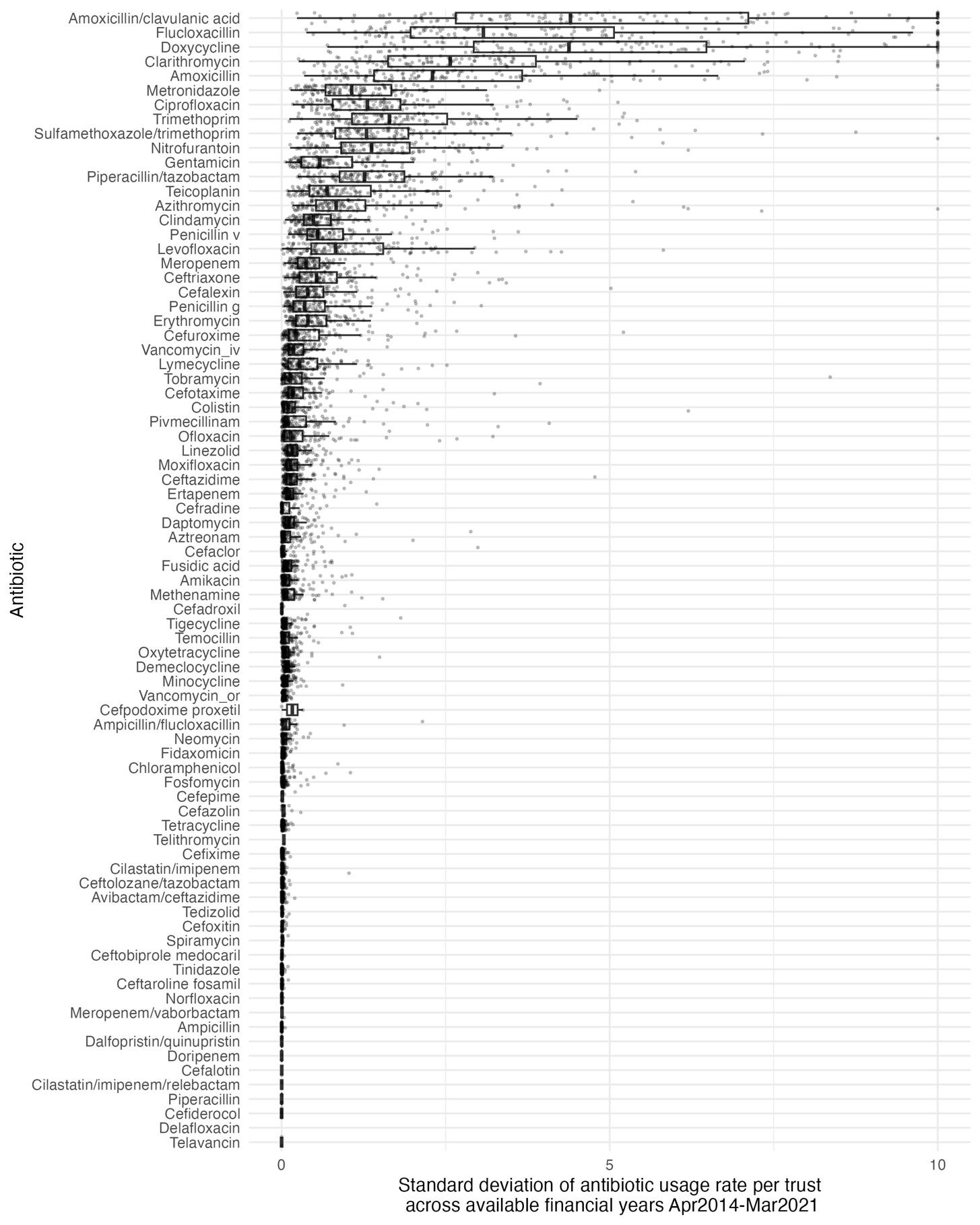

Note: Outliers outside of x-axis scale (absolute value >10) were truncated.

Figure S6 Distribution of antibiotic usage per antibiotic across Trusts and financial years, for the top 24 most used antibiotics.

(A)

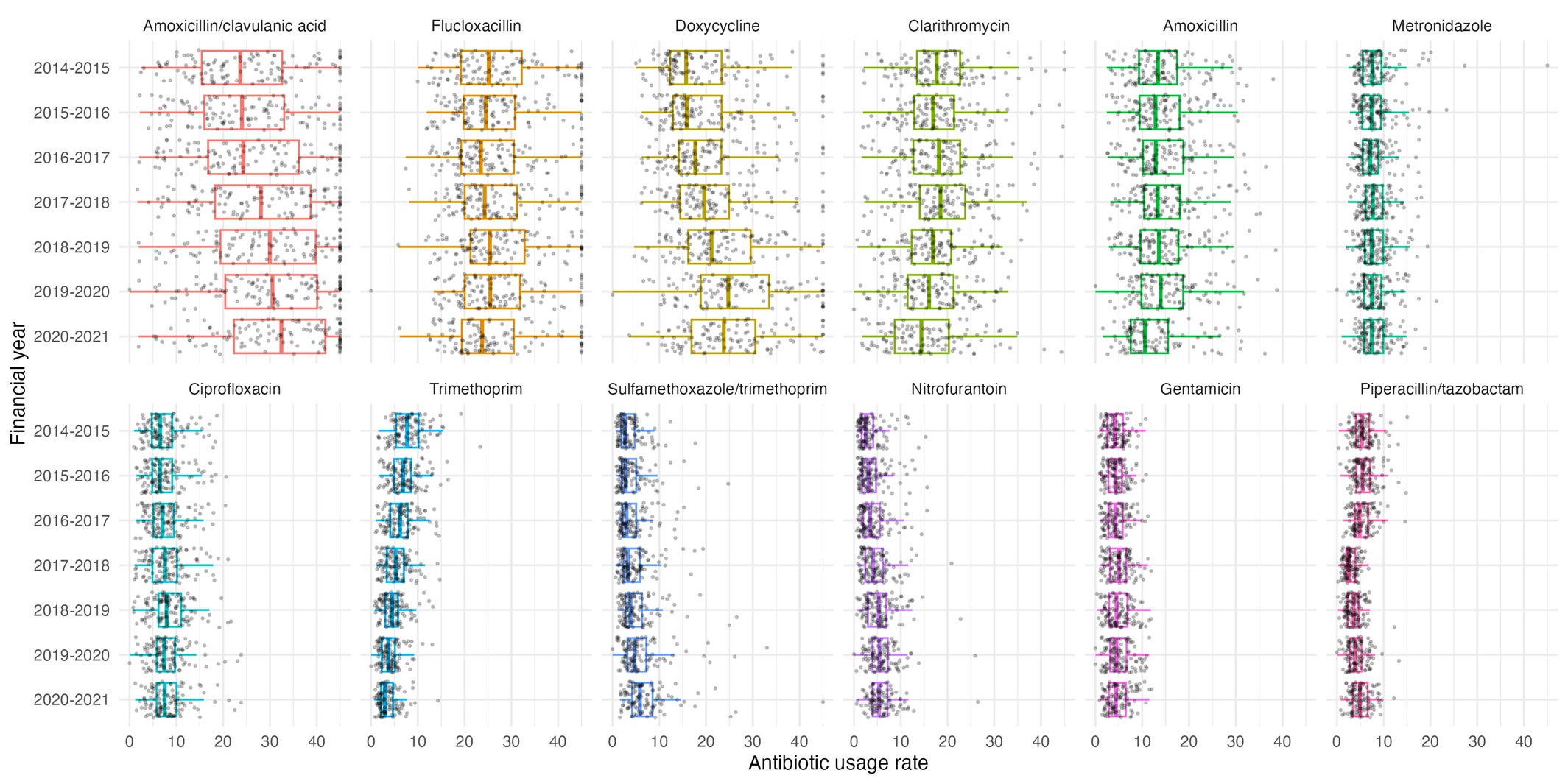

(B)
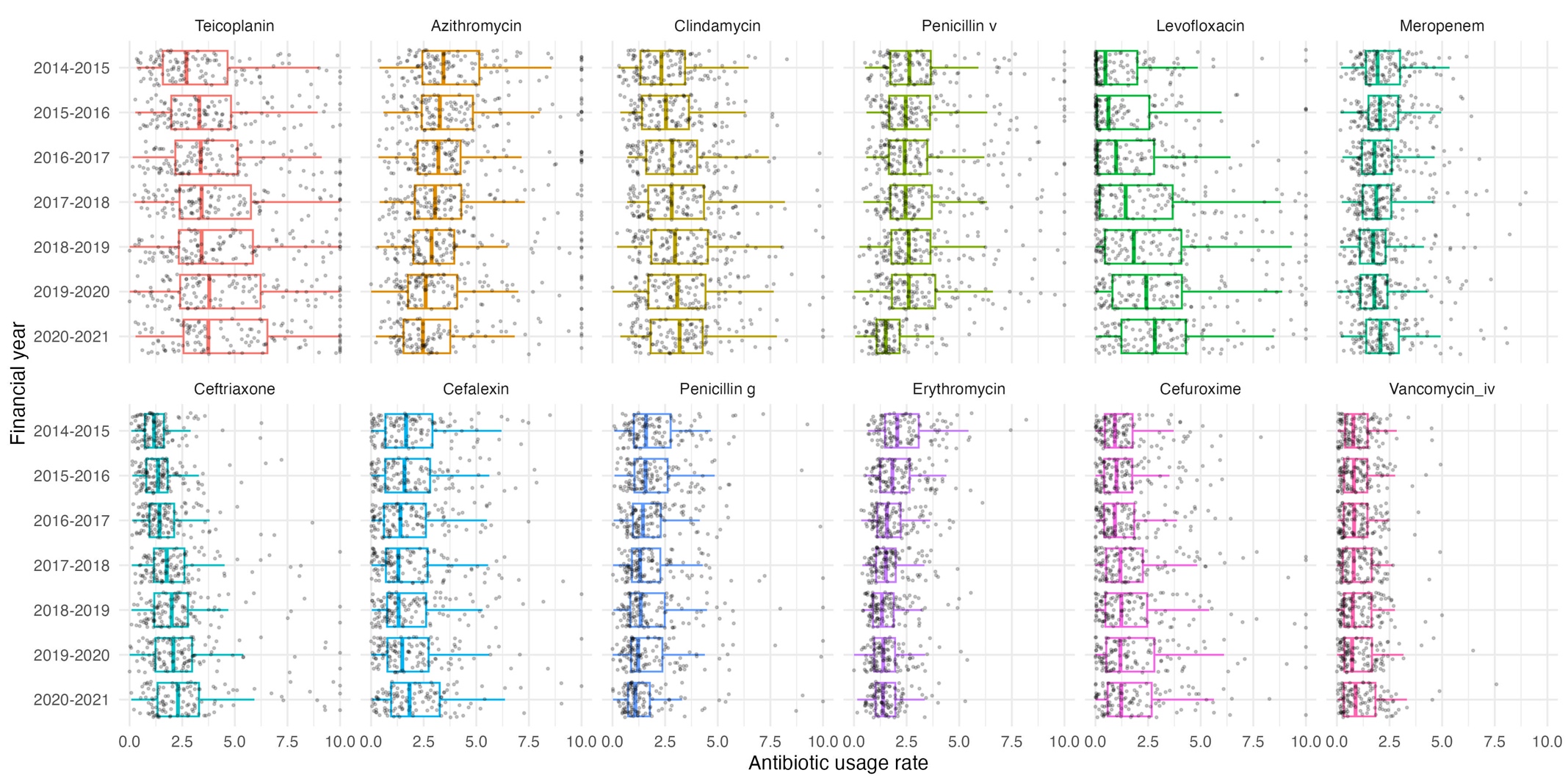

Note: Panel (A) and (B) have different scales on the y-axis. Outliers outside of x-axis scale (absolute value >10) were truncated.

Figure S7 Distribution of difference between the current and previous financial year antibiotic usage rate per antibiotic across all Trust-FYs for the top 12 most used antibiotics.

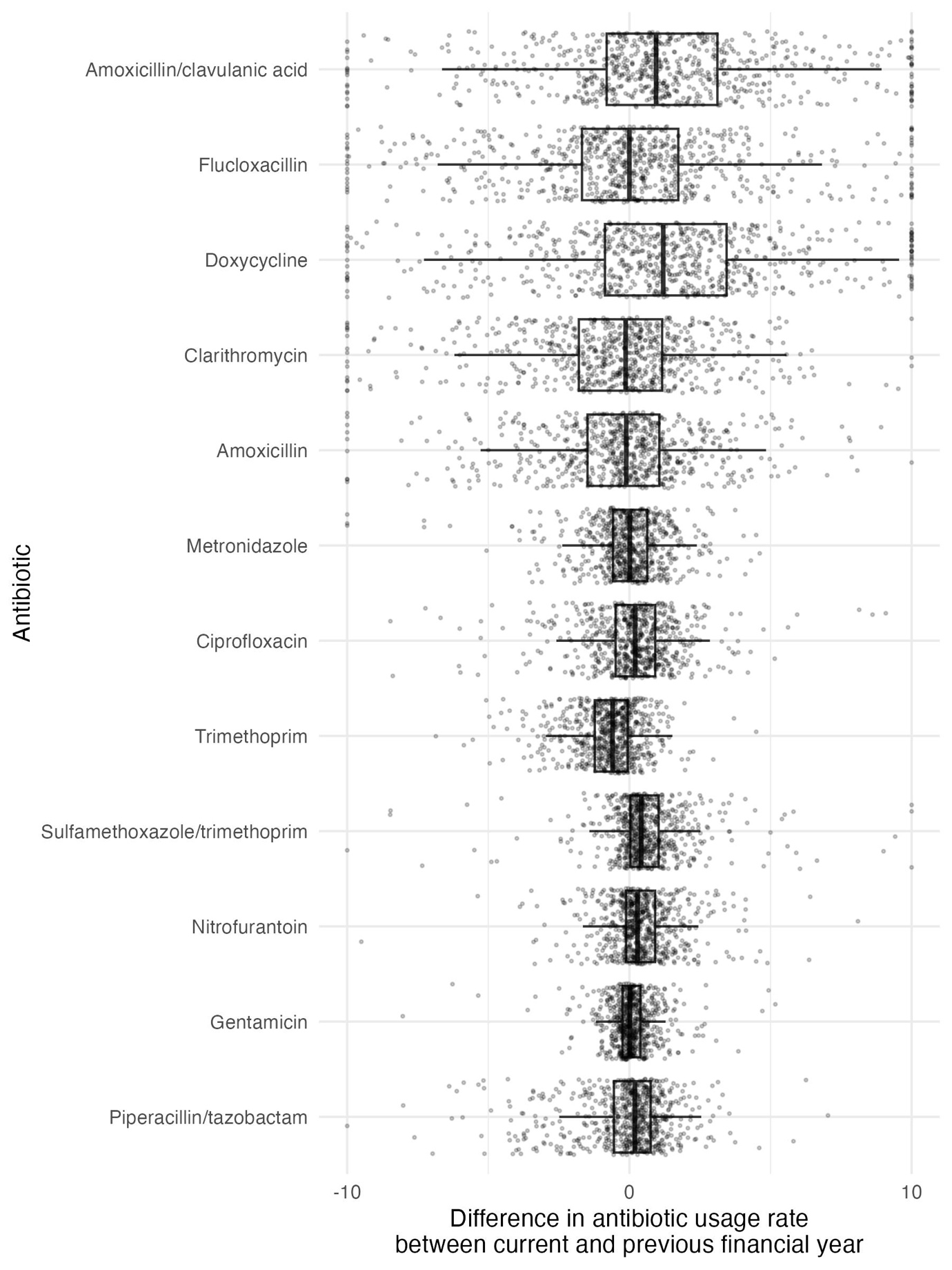

Note: one point per Trust-antibiotic-FY. Outliers outside of x-axis scale (absolute value >10) were truncated.

Figure S8 Distribution of difference between the current and previous financial year antibiotic usage rate by financial years across all trusts for the top 12 most used antibiotics.

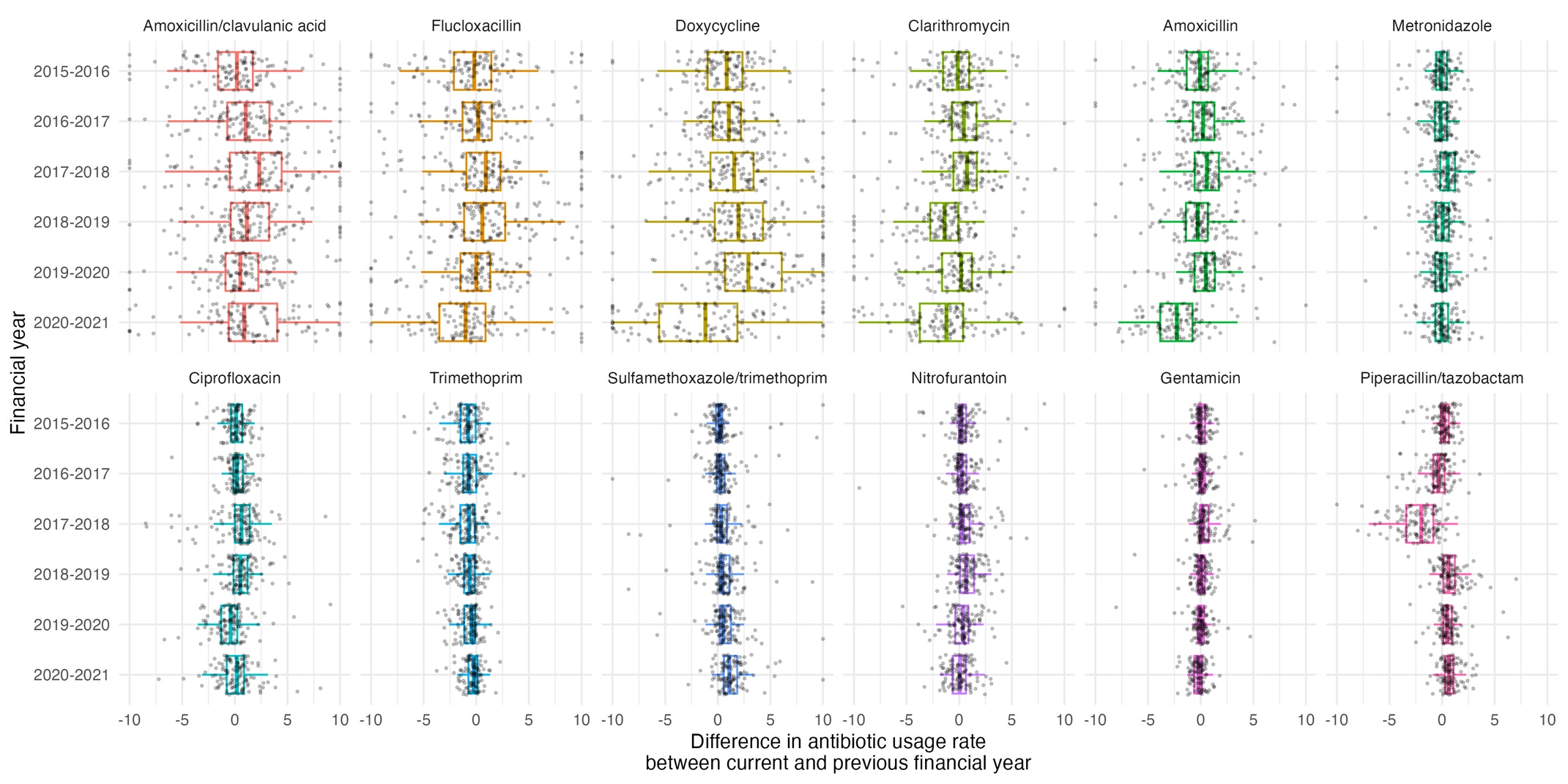

Note: one point per Trust-antibiotic. Note: Outliers outside of x-axis scale (absolute value >10) were truncated.

Figure S9 Mean absolute error of predicting current resistance prevalence from previous value taken forwards by pathogen-antibiotic-FY over Trusts.

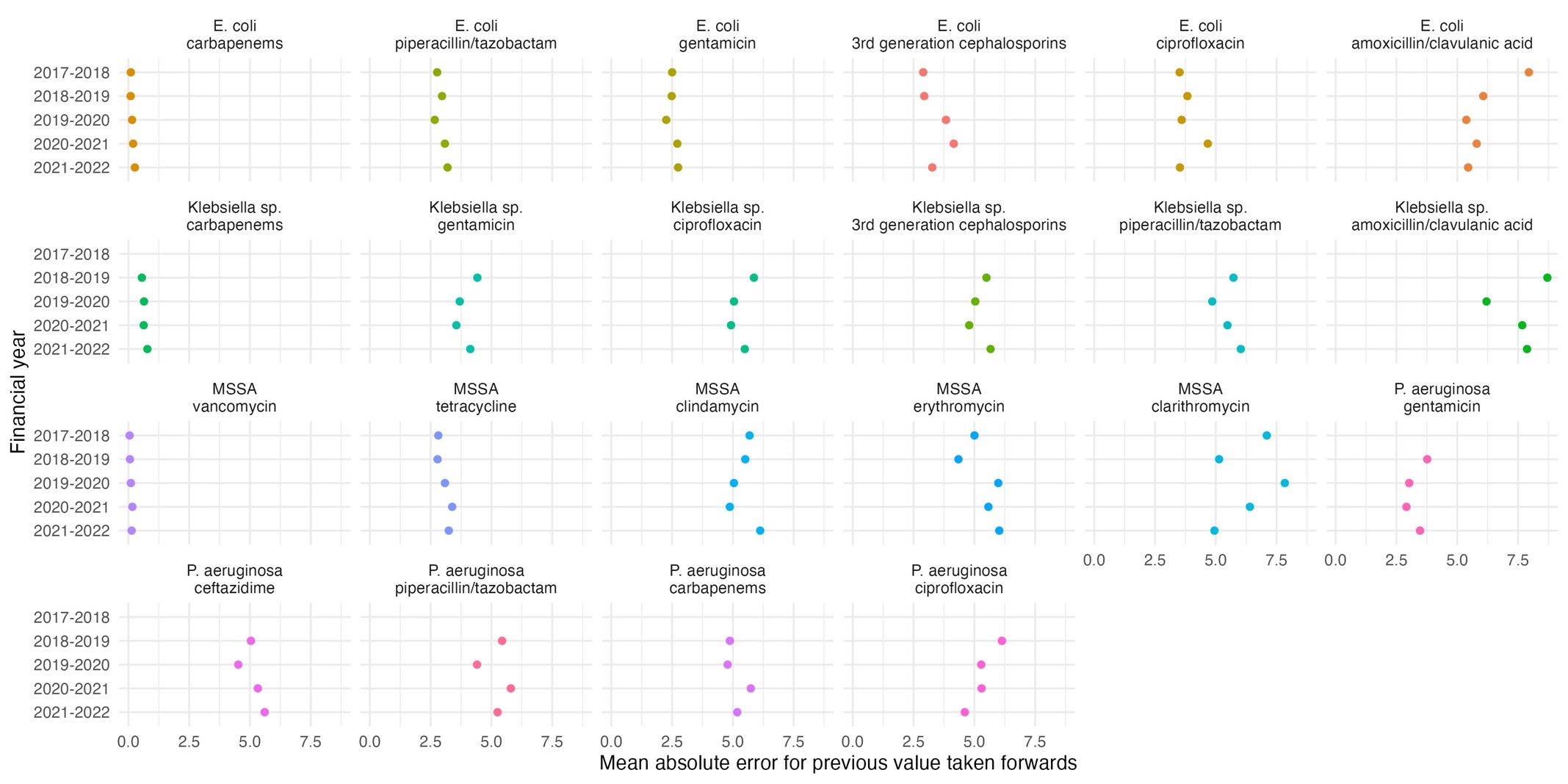

Figure S10 Mean absolute error for prediction on test set (percentage resistance in FY2021-2022) for 6 different XGBoost prediction models: considering antibiotic usage in the previous year alone as input features (no information on previous resistance prevalence) and increasing the size of the training dataset by considering previous years as additional outcomes (XGBoost usage (1yr, “1yr double” and “1yr triple”)), as well as XGBoost models with 3, 2 and 1 FY(s) historical data, for both usage and resistance (increasing the size of the training dataset by considering previous years as additional outcomes)

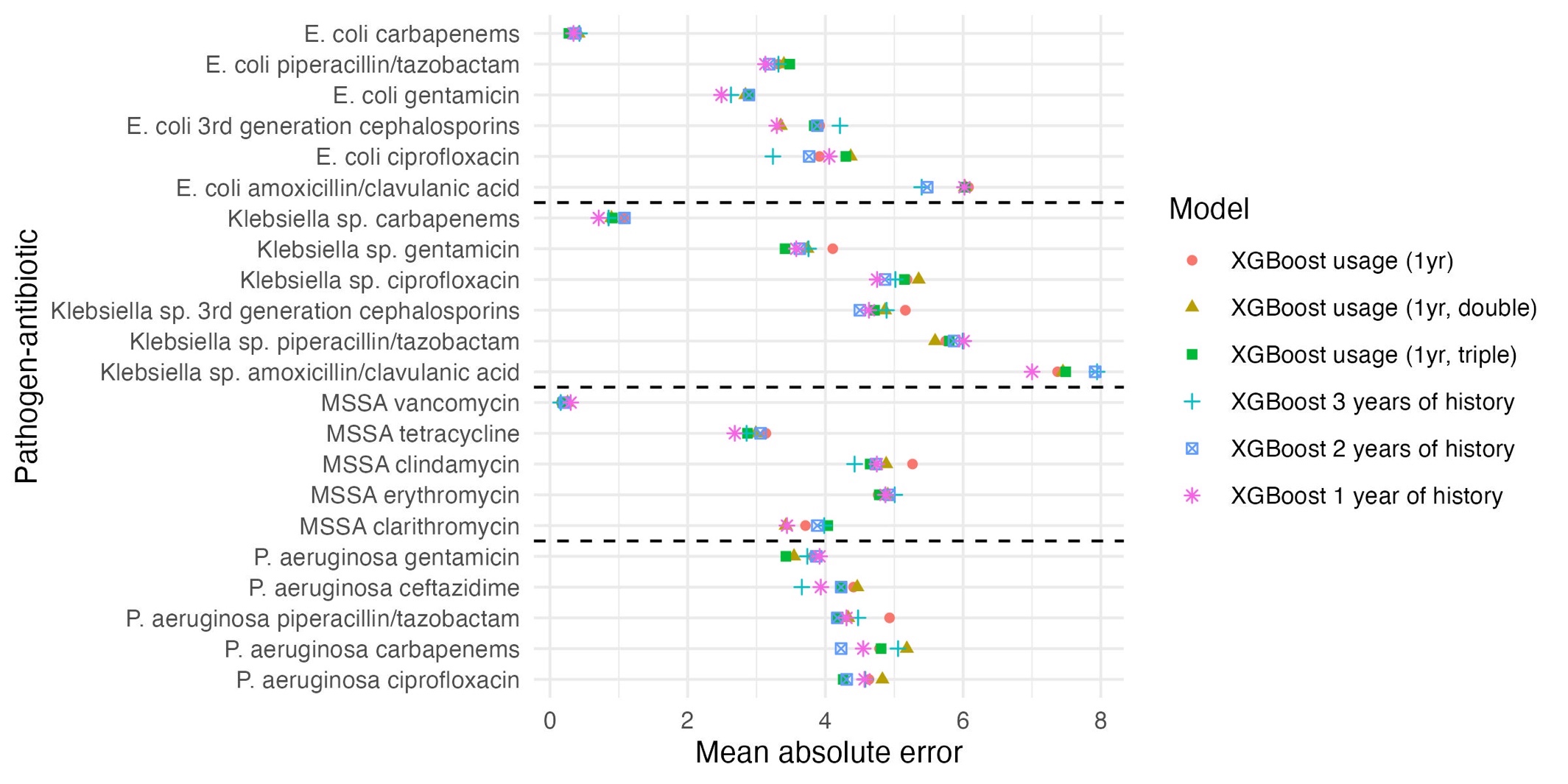

Figure S11 Mean absolute error for prediction on test set (percentage resistance in FY2021-2022) for previous value taken forwards and 2 different XGBoost prediction models: XGBoost with all historical usage and resistance prevalence (XGBoost default), and XGBoost with selected features based on ranking according to mean absolute SHAP values being above that of an additional feature representing white noise (XGBoost default SHAP fs)

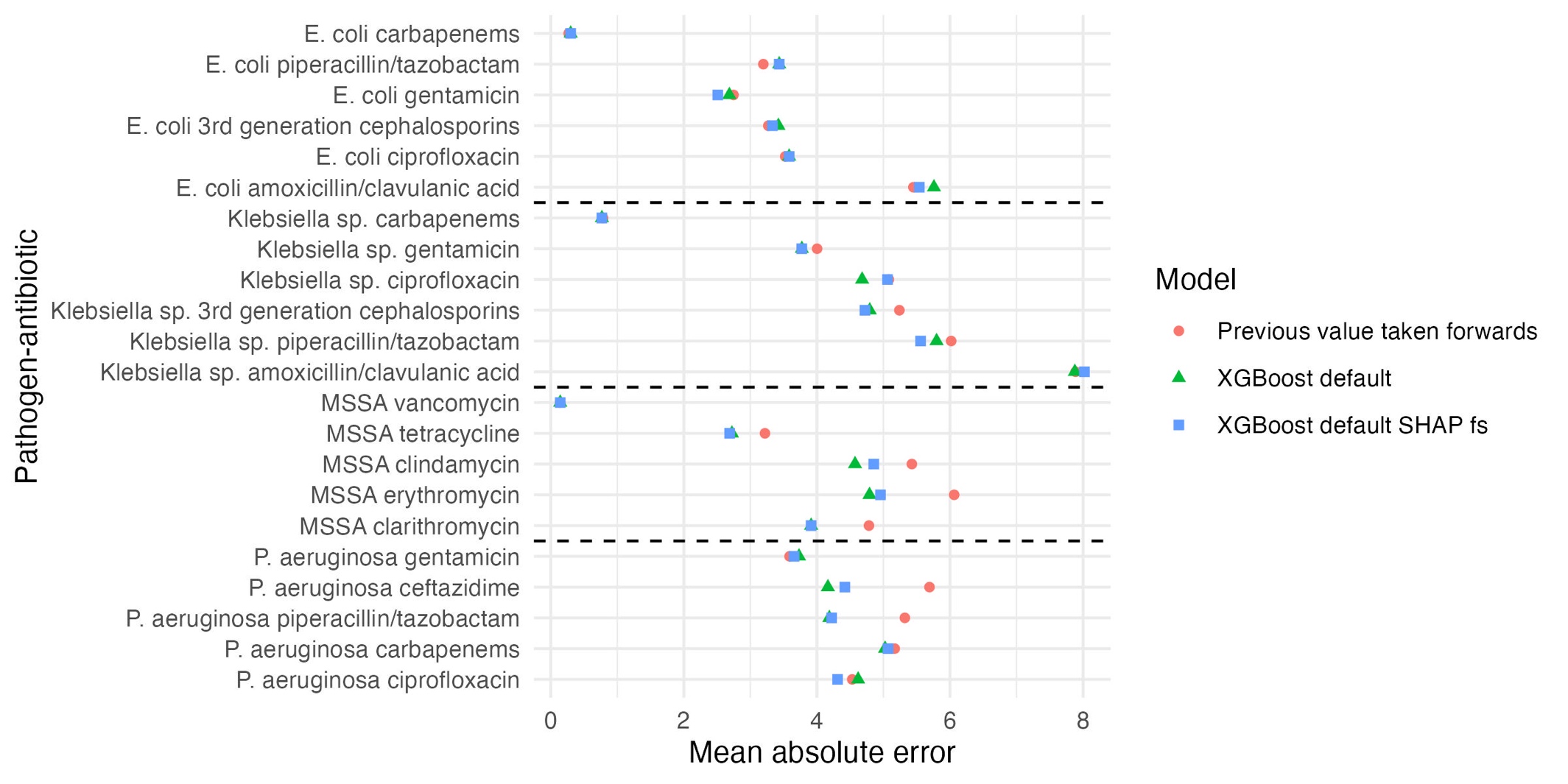

Figure S12 Mean absolute error for prediction on test set (resistance prevalence in FY2021-2022) for 6 different prediction models split by absolute difference between FY2021-2022 and FY2020-2021 resistance prevalence >7.5% or ≤7.5% (A), >5% or ≤5% (B)

(A)

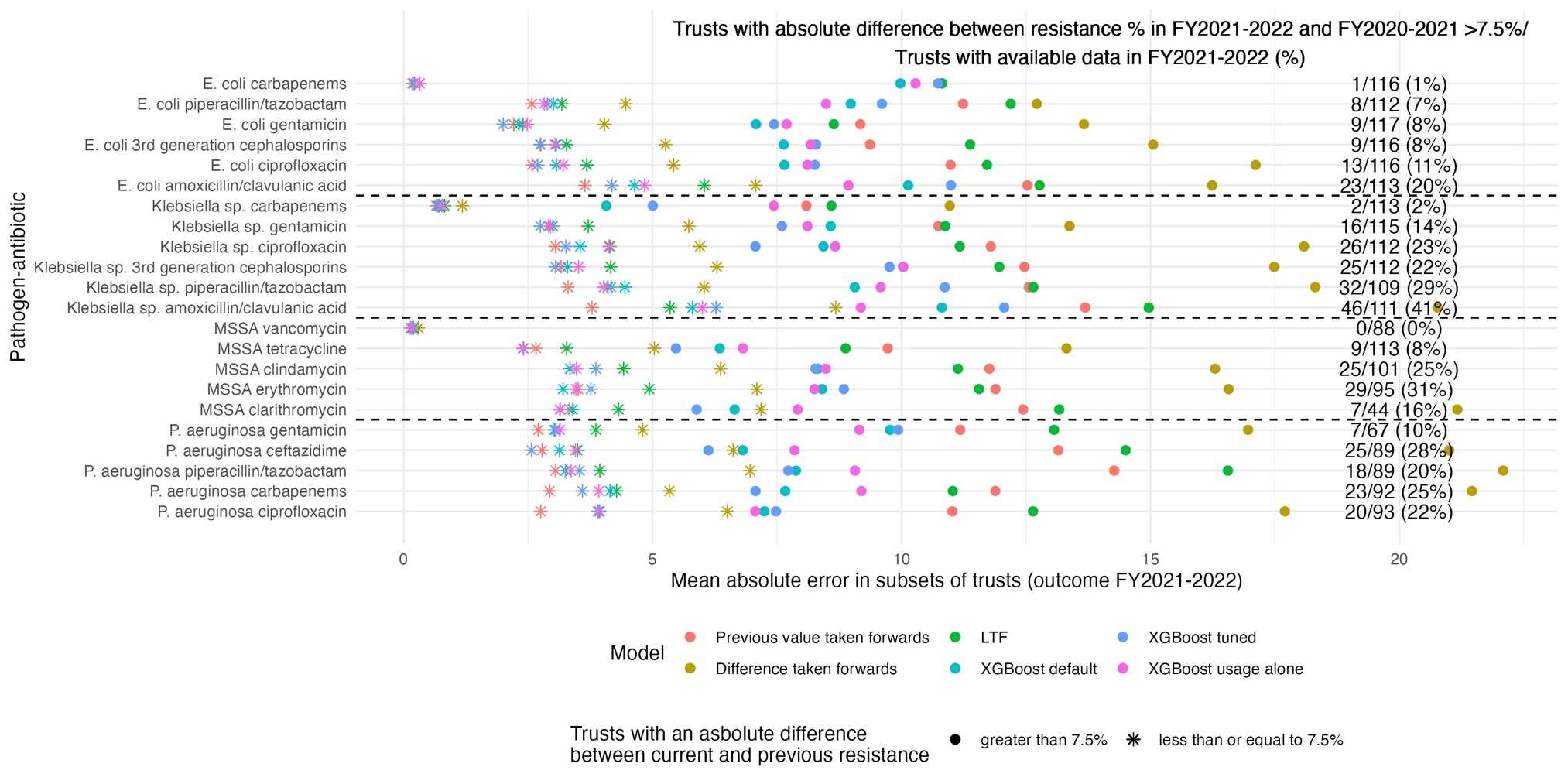

(B)

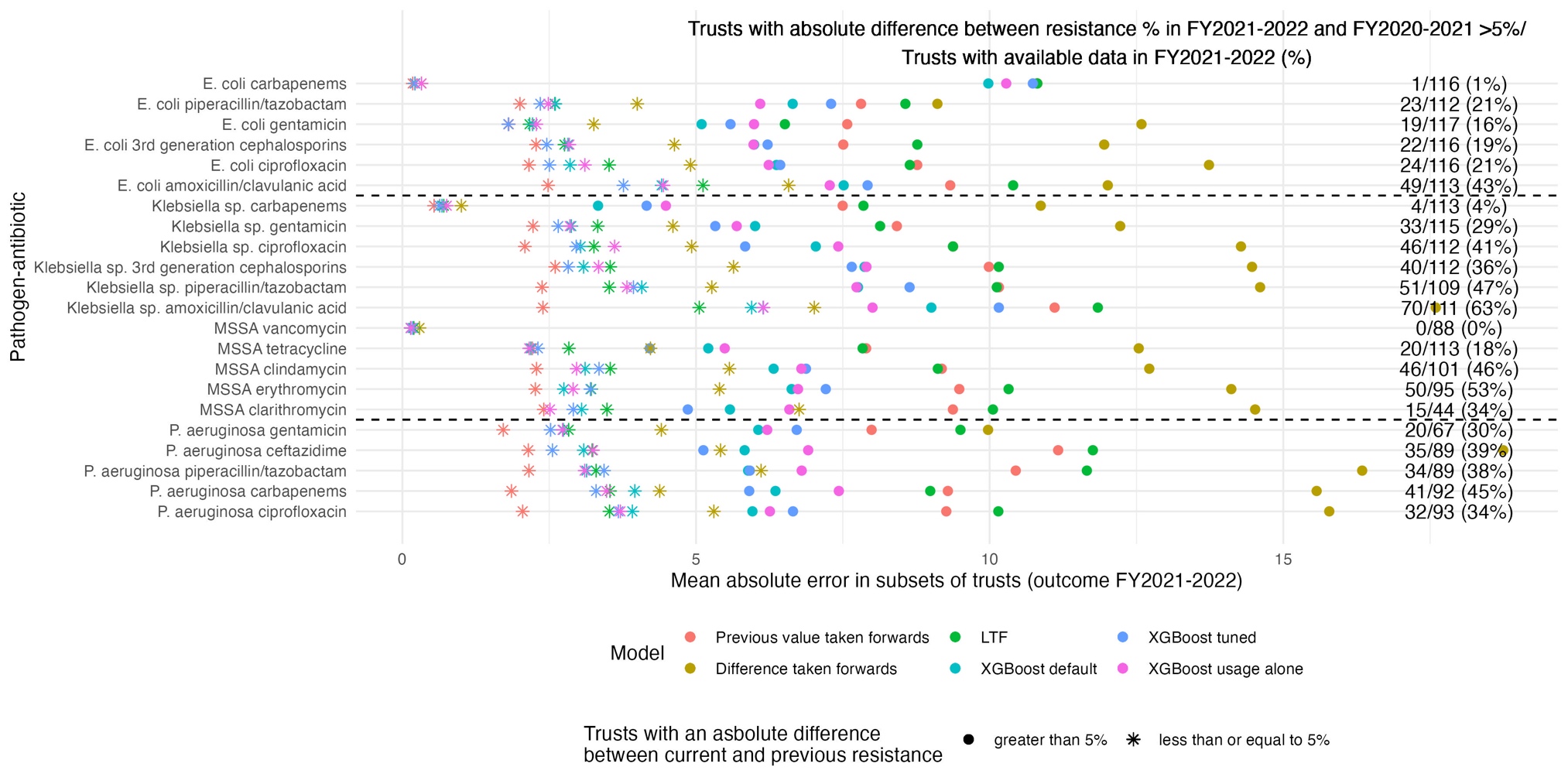

Note: 70 residuals that had either missing previous value or previous difference were excluded for comparability of performance measures between the models, although XGboost also made these predictions.

Figure S13 Mean absolute error for prediction on test set (resistance prevalence in FY2021-2022) for 6 different prediction models in those Trusts with an absolute difference between FY2021-2022 and FY2020-2021 resistance prevalence >10%, split by whether the current resistance prevalence was greater or lower than the previous resistance prevalence

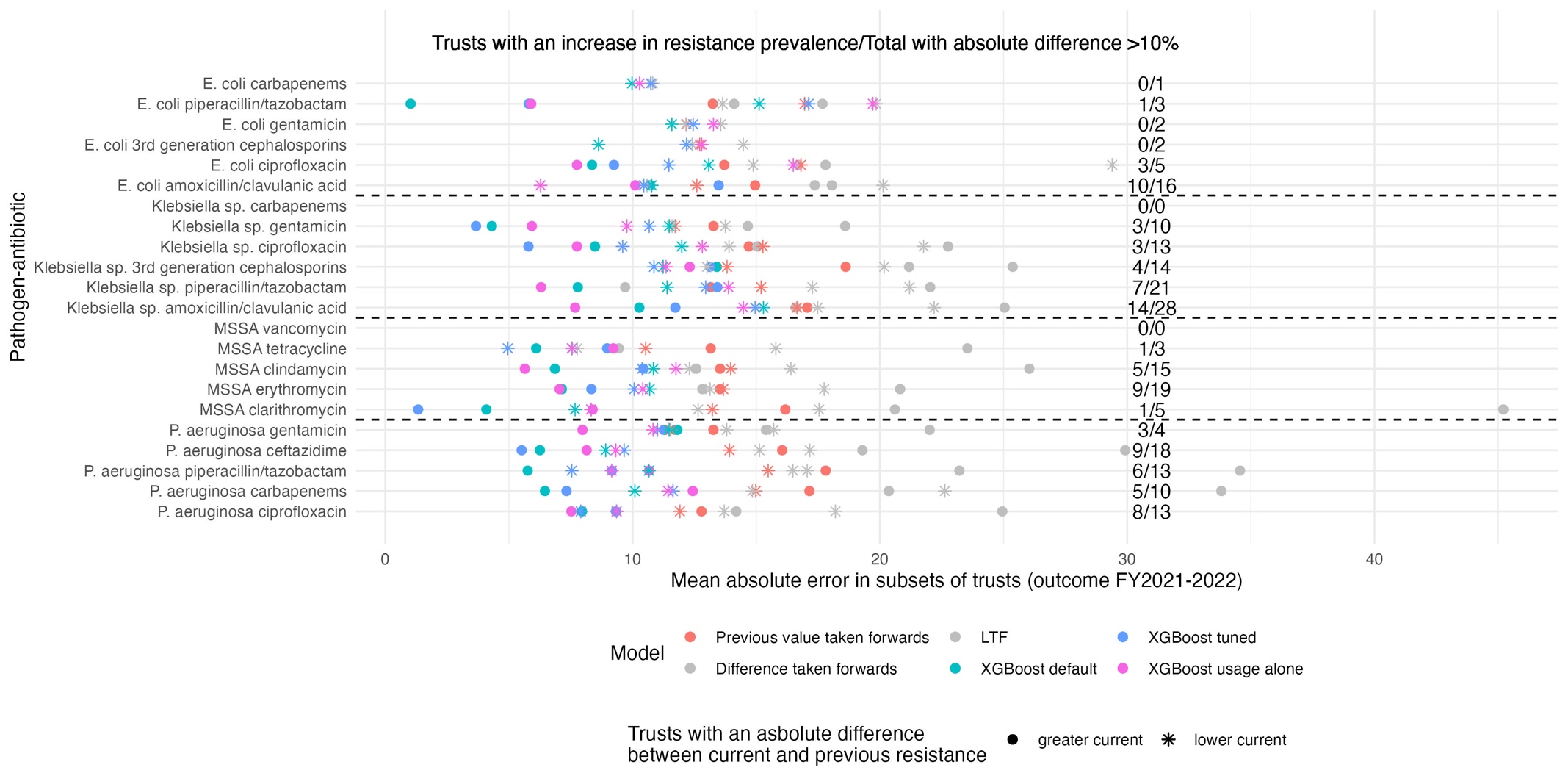
